# The polygenic architecture of hidradenitis suppurativa reveals signaling mechanisms that implicate epithelial remodeling

**DOI:** 10.1101/2025.07.25.25332168

**Authors:** Atlas Khan, Poppy A. Gould, Yiming Luo, Errol P. Prens, Lee Wheless, Adriana M. Hung, VA Million Veteran Program, Theodore G. Drivas, Marylyn D. Ritchie, Amir Hossein Saeidian, Hákon Hákonarson, Michael March, Nick Dand, Jonathan Barker, Michael Simpson, Jake Saklatvala, Xinyi Du-Harpur, Shahir Farnood, Raymond Chung, Charles J Curtis, Sang Hyuck Lee, Brian Kirby, Maris Teder- Laving, Külli Kingo, Estonian Biobank research team, Laurent F. Thomas, Mari Løset, Ben Michael Brumpton, Kristian Hveem, M. Geoffrey Hayes, John Connolly, Frank Mentch, Patrick Sleiman, Kathleen LaRow Brown, Nicholas Tatonetti, Olivia D. Perez, Alice Braun, Stephan Ripke, Sadhana Gaddam, Anthony Oro, Leah C. Redmond, Claire Higgins, Meng-Ju Lin, Ernest S. Chiu, Catherine P. Lu, George Hripcsak, Chunhua Weng, Krzysztof Kiryluk, Lam C. Tsoi, Johann E. Gudjonsson, Kelsey R. van Straalen, Joshua D. Milner, Lynn Petukhova

## Abstract

We sought to identify clinically relevant regulators of hair follicle inflammation by conducting a human genetic study of hidradenitis suppurativa (HS), a prevalent, understudied, inflammatory disease with limited effective treatments. We performed a GWAS with 6,300 cases and identified 12 independent risk loci. Epigenetic and transcriptomic analyses of HS risk variants defined cell-specific gene regulatory programs. We experimentally validated a coherent gene module defined by upregulated *SOX9*, *CXCR4*, and *CD74* co-expression that maps to aberrant epithelial structures in the skin. Pharmacological inhibition of CXCR4 implicates CD74 mediated regulation of PI3K/AKT and NF-κB signaling to calibrate inflammation, proliferation and apoptosis in keratinocytes. We next used genome-wide methods to interrogate shared polygenic architecture and identified new clinically and mechanistically relevant disease associations, including another condition that involves aberrant hair follicle remodeling, male pattern hair loss. Our results point towards CXCR4-CD74 signaling in HS and hair follicle homeostasis and suggest CXCR4 blockade as a new therapeutic strategy in HS.

## Main Text

Human genetic studies identify key physiological regulators while also providing insight into the clinical effects of perturbing them. Genetic studies allow us to use human diseases to discover and model biological mechanisms of development and homeostasis that have direct relevance to human biology and medicine.

The hair follicle is an ideal model for understanding fundamental principles of human tissue regeneration (*1*). It undergoes physiological remodeling as it cycles through periods of growth, regression, and rest. It also contributes to wound-induced cutaneous remodeling through stem cell fate switching (*2*). Appropriately regulated inflammation remains crucial in both contexts, governing stem cell activation, proliferation, migration, and differentiation (*3, 4*).

Hidradenitis suppurativa (HS) is a prevalent but understudied inflammatory skin disease. Despite affecting 1% of the population, there are only three FDA approved treatments, and these fail to provide sustained benefits for most patients. HS has a distinctive clinical presentation, with inflammation occurring at terminal hair follicles in intertriginous skin (*5*). Unmitigated epithelial proliferation in the hair follicle causes follicular infundibular hyperplasia that leads to hair follicle occlusion, distension and rupturing. Histological hallmarks of HS also include epithelial tendrils that extend downward into the dermis and epithelialized dermal tunnels. HS shares clinical features of pathological inflammation with a broader class of inflammatory diseases that affect diverse organs, including impaired healing, scarring that causes permanent destruction of tissue architecture, and persistent, debilitating pain. Controversy exists over the cellular origins and initiating and exacerbating factors that drive this common and difficult to manage disease.

The genetic architecture of HS has been insufficiently studied despite an urgent need for mechanistic, genetically informed insights to guide novel therapeutic strategies. There is robust statistical evidence for cosegregation of rare loss of function (LOF) variants in γ-secretase genes, *NCSTN* and *PSENEN* (*6*). While the mechanism remains to be fully understood, downstream biological effects of γ-secretase LOF include attenuated Notch signaling and upregulated PI3K/Akt signaling (*7*). There is evidence that more single-gene causes of HS await discovery and that these will reveal etiological heterogeneity requiring precision medicine approaches to disease management (*7*). Two HS genome-wide association studies (GWAS) meta-analyses have been reported, which together have identified common variant associations at eight loci (*8, 9*). Three loci contain population-specific low frequency protein coding variants in *NCSTN*, *PSENEN*, and *WNT10A* respectively. At the *WNT10A* locus, a missense coding variant protective of HS was previously known to cause hair follicle miniaturization in children and adults (*10*). Variants at the remaining five loci are intergenic. Of these, the strongest effects are seen at two loci that are reported in both studies, chromosomes 17q24.3 and 13q22.1. Variants at these loci localize most closely to *SOX9* and *KLF5* respectively – both transcription factors (TF) with well-established roles in hair follicle development and inflammation. They are also each linked to both NOTCH and PI3K/AKT signaling (*11, 12*). Risk loci are also reported in the *HLA* locus and at 9q31.3 and 14q24.3. Genetic and disease mechanisms driving these variant associations remain largely unexplored.

Here, we expand upon our knowledge of the polygenic architecture of HS by conducting a genome-wide association study in an ancestrally complex cohort of 6,361 cases and 1,520,765 controls, investigating the regulatory mechanisms that coordinate gene expression across loci, and characterizing biological and clinical effects of HS risk alleles, which lend to rational targets for precision intervention.

## Results

### Genome-wide association studies identify 12 pan-ancestry and two ancestry-specific loci

We conducted a meta-analysis GWAS across 10 cohorts, aggregating data from a total of 1,527,126 subjects, which included 6,361 cases and 1,520,765 controls. The characteristics of the subjects in each cohort are summarized in **Table 1** and described in further detail in the **Supplementary Note**.

**Table 1.** Cohort Details for Multi-Ancestry HS GWAS.

|  | All |  | EUR cohort |  | AFR cohort |  | AMR cohort |  |
| --- | --- | --- | --- | --- | --- | --- | --- | --- |
| Source | Cases | Controls | Cases | Controls | Cases | Controls | Cases | Controls |
| eMERGE-III | 544 | 80,323 | 302 | 66522 | 242 | 13,801 | - | - |
| University College Dublin | 223 | 7,072 | 223 | 7,072 | - | - | - | - |
| CHOP-GSA array | 130 | 32,226 | 26 | 17,026 | 104 | 15,200 | - | - |
| CHOP-HM | 58 | 10,582 | 15 | 5,785 | 43 | 4,797 | - | - |
| Michigan | 396 | 47,980 | 290 | 44,964 | 106 | 3,016 | - | - |
| Penn | 275 | 32,492 | 67 | 25,055 | 208 | 7,437 | - | - |
| Estonianbio bank | 334 | 200,653 | 334 | 200,653 | - | - | - | - |
| MVP | 2,348 | 575,878 | 1,271 | 455,878 | 1,077 | 119,700 |  |  |
| AoU | 888 | 118,326 | 407 | 73,129 | 333 | 21,950 | 148 | 19,611 |
| FinGen | 800 | 328747 | 800 | 328747 | - | - | - | - |
| Hunt | 365 | 86486 | 365 | 86486 | - | - | - | - |
| Total | 6,361 | 1,520,765 | 4,100 | 1,311,317 | 2,113 | 185,901 | 148 | 19,611 |

Our primary analysis was a combined cohort meta-analysis that identified 126 variants at 12 loci with genome-wide significance (p< 5.0E-08) and minimal evidence for genomic inflation (lambda=1.006) (**Figure 1**). The regional plots for each locus are shown in **Supplementary Figure 1**. This analysis replicated four loci previously associated with HS (6p21.32; the HLA, 9q31.3, 13q22.1, and 17q24.3) (*8, 9*). We additionally discovered 8 new genome-wide significant loci, including a second independent risk haplotype at chromosome 13q22. (**Table 2**). Stepwise conditional analyses also identified multiple independent associations at the *HLA* locus. None of the significant variants alter the sequence or structure of proteins. Protein-coding transcripts are carried on six risk haplotypes, including *CYP1B1*, *CXCR4*, *KLF5*, *RBBP8*, *MAPRE2*, and *KREMEN1* respectively.

**Figure 1.**
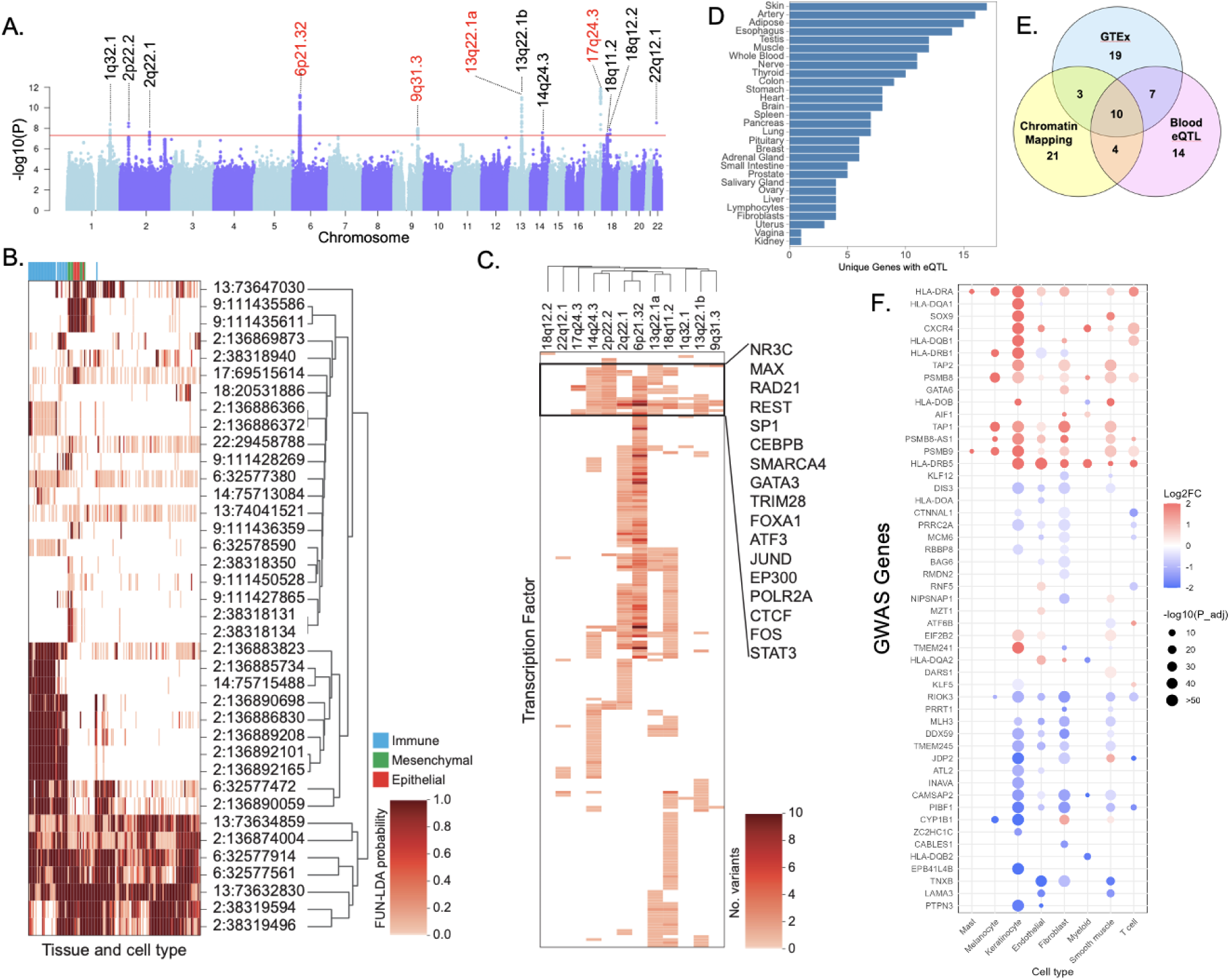
Mapping HS risk variants to cis regulatory elements and genes. (A) Meta-analysis GWAS identified 12 independent associations including 4 previously reported loci (red) and 8 new loci (black). (B) Hierarchical clustering of HS risk variants predicted to have functional effects in Roadmap and Encode samples identifies a cluster of variants in regions that are predicted to be active in immune cells and tissues (blue) and a cluster of variants predicted to be active in epithelial (red) and mesenchymal derived or derived mesenchymal (green) cells. (C) HS risk variants fall within transcription factor (TF) bind sites that are shared across GWAS loci. (D) HS risk variants were mapped to eQTL data in GTEx. Skin is the most highly represented tissue. (E) HS risk variants were linked to 79 transcripts through eQTL or chromatin mapping data. Each data set implicates overlapping sets of genes. (F) Genes predicted to be influenced by HS risk variants were tested for differential gene expression in single cell transcriptomic data comparing HS lesional skin to normal healthy skin.

**Table 2.** HS GWAS Genome Wide Significant Loci by Ancestry Cohort.

| Region | CHR | BP (Hg38) | rs ID | Alt/Ref | EUR cohort<br>OR 95%<br>CI, P | AFR cohort<br>OR 95%<br>CI, P | AMR cohort<br>OR 95%<br>CI, P | Meta<br>Analysis<br>OR 95%<br>CI, P |
| --- | --- | --- | --- | --- | --- | --- | --- | --- |
| 1q32.1 | 1 | 200918529 | rs296539 | T/C | 1.12 (1.07-1.17), | 1.11 (1.03-1.19), | 1.02 (0.80- | 1.11 (1.07-1.16), |
|  |  |  |  |  | P=8.82E-07 | P=3.93E-03 | 1.31),<br>P=0.85 | P=1.44E-08 |
| **2q22.1** | 2 | 136128802 | rs7588118 | C/G | 1.11 (1.06-1.18),<br>P=1.01E-04 | 1.17 (1.09-1.25),<br>P=2.68E-05 | 0.94 (0.71-1.24),<br>P=0.65 | 1.13 (1.08-1.18),<br>P=3.90E-08 |
| **2p22.2** | 2 | 38092354 | rs162330 | A/C | 1.13 (1.08-1.18),<br>P=9.24E-08 | 1.10 (1.02-1.19),<br>P=1.68E-02 | 1.14 (0.88-1.46),<br>P=0.32 | 1.12 (1.08-1.16),<br>P=3.15E-09 |
| **6p21.32** | 6 | 32614196 | rs521539 | A/G | 1.19 (1.13-1.26),<br>P=3.37E-10 | 1.13 (1.02-1.24),<br>P=1.55E-02 | 1.22 (0.93-1.61),<br>P=0.15 | 1.18 (1.12-1.23),<br>P=9.90E-12 |
| **9q31.3** | 9 | 108688248 | rs10816700 | A/G | 1.15 (1.10-1.20),<br>P=8.62E-10 | 1.02 (0.95-1.11),<br>P=0.58 | 1.15 (0.90-1.46),<br>P=0.27 | 1.12 (1.07-1.16),<br>P=1.03E-08 |
| **13q22.1a** | 13 | 73060721 | rs41286076 | T/C | 0.84 (0.80-0.89),<br>P=7.09E-10 | 0.87 (0.75-1.00),<br>P=5.40E-02 | 1.05 (0.76-1.44),<br>P=0.78 | 0.85 (0.81-0.89),<br>P=2.32E-10 |
| **13q22.1b** | 13 | 73473464 | rs9530199 | T/G | 0.81 (0.76-0.86),<br>P=6.36E-12 | 0.97 (0.83-1.12),<br>P=0.65 | 0.63 (0.43-0.93),<br>P=0.01 | 0.82 (0.78-0.87),<br>P=1.01E-11 |
| **14q24.3** | 14 | 75242292 | rs8011988 | A/G | 0.88 (0.84-0.92),<br>P=3.34E-08 | 0.92 (0.84-1.02),<br>P=0.1046 | 1.04 (0.81-1.35),<br>P=0.74 | 0.89 (0.86-0.93),<br>P=2.65E-08 |
| **17q24.3** | 17 | 71514343 | rs73994811 | T/G | 1.22 (1.14-1.30),<br>P=9.48E-09 | 1.18 (1.09-1.28),<br>P=4.85E-05 | 1.10 (0.83-1.45),<br>P=0.50 | 1.20 (1.14-1.26),<br>P=2.42E-12 |
| **18q11.2** | 18 | 23006282 | rs72889626 | T/C | 0.85 (0.81-0.90),<br>P=1.12E-08 | 0.96 (0.83-1.12),<br>P=0.64 | 0.98 (0.73-1.31),<br>P=0.88 | 0.87 (0.82-0.91),<br>P=4.05E-08 |
| **18q12.2** | 18 | 35198170 | rs118127088 | A/G | 1.53 (1.32-1.77),<br>P=1.56E-08 | 1.29 (0.46-3.60),<br>P=0.62 | 0.77 (0.31-1.93),<br>P=0.57 | 1.50 (1.30-1.73),<br>P=3.57E-08 |
| **22q12.1** | 22 | 29062800 | rs16987014 | A/G | 0.77 (0.70-0.85),<br>P=9.37E-08 | 0.77 (0.65-0.92),<br>P=4.05E-03 | 1.10 (0.64-1.90),<br>P=0.74 | 0.78 (0.71-0.84),<br>P=2.90E-09 |
| **Replication of Andersen et al., 2025** |  |  |  |  |  |  |  |  |
| **2q35** | 2 | 218890289 | rs121908120 | T/A | 1.66 (1.23–2.24),<br>P=1.05E-03 | - | - | 1.66 (1.23–2.24),<br>P=1.05E-03 |
| **14q24.3** | 14 | 75230213 | rs17103088 | G/A | 1.15 (1.09–1.21), | 1.10 (0.92–1.32), | 1.13 (0.83–1.55), | 1.14 (1.09–1.20), |
|  |  |  |  |  | P=5.34E-07 | P=0.283 | P=0.444 | P=2.35E-07 |
| **1q23.2** | 1 | 160354223 | rs771414318 | C | - | - | - | - |
| **19q13.12** | 19 | 35746829 | - | A | - | - | - | - |

We next performed ancestry-specific meta-analyses. In the European ancestry cohort we identified one additional significantly associated locus at chromosome 5p13.2 (rs4242248-T, OR=0.88 95%, CI:0.84-0.92, P=4.08E^-08^) (**Supplementary Figure 2**). In the African American cohort we identified an additional associated locus at 4p12 (rs10461071-A OR=1.50, 95% CI:1.31-1.73, P=5.35E^-09^) (**Supplementary Figure 3**). At both loci, allele frequencies and LD patterns differ between European and African ancestral groups and each of the ancestry-specific risk haplotypes was found to carry population-specific low-frequency missense variants (D′=1). In European ancestry populations, rs4242248 is correlated most strongly with rs61734063 *RANBP3L* p.Thr92Pro (D′=1), and also has strong correlations (D′>0.9) with variants in *IL7*, *SPEF2*, and *NADK2*. Lower correlations are found in African populations (D’<0.45). In African ancestry populations, rs10461071 is correlated with rs565358308 *GUF1* p.Glu653Val. (D′=1), a variant that is not present in European populations.

Overall, our meta-analysis identified 12 pan-ancestry risk loci that contain 126 genome-wide significant variants. To investigate genetic and epigenetic effects mediating these associations, we also included an additional 88 variants that are in high LD (r2>.8) with index variants, as reported by the 1000 Genomes Project (**Supplementary Data 1**)(*13*).

### Shared gene regulatory mechanisms span independent HS-risk loci

Since most GWAS variants contribute to disease risk by perturbing cis-regulatory elements (cREs), we mapped HS risk variants to tissue-specific active regulatory elements. We utilized FUN-LDA (*14*), a model trained on data from the Roadmap Epigenomics Project – specifically active histone marks (H3K4me1, H3K4me3, H3K9ac, and H3K27ac) and DNase Hypersensitivity Sites (HSS). We identified 38 HS risk variants across 10 loci have a high posterior probability (PP>.7) of having tissue-specific active regulatory activity (**Supplementary Data 2; Supplementary Figures 4-13**). Hierarchical clustering of variants and tissues identified three major clades (**Figure 1B**). One clade includes variants in active regions across most tissues, likely indicating ubiquitously active regulatory elements. In contrast, the second and third clades are indicative regulatory activity in cells and tissues that are relevant to HS. The second clade is enriched for immune cells and tissues (e.g., hematopoietic stem cells (HSCs), B-cells, T-cells, and thymus), while the third clade includes epithelial cells, mesenchymal stem cell (MSC)-derived tissues, and tissue-derived MSCs. The combination of cells and tissues in this third clade is reminiscent of an epithelial-to-mesenchymal transition (EMT) phenotype, as is seen in keratinocytes that become activated during re-epithelialization of cutaneous wounds (*15*). Given the sex disparities in HS (*16*), it is interesting to note that our analysis also highlighted sex-specific regulatory activity: HS variants at loci on chromosomes 2, 6, and 22 fell within cREs that are active in female but not male skeletal muscle; while variants on chromosomes 6, 9, and 13 showed male specific activity in mobilized HSCs.

GWAS variants in cREs can also perturb transcription factor binding thereby disrupting gene expression networks (*17*). To investigate this, we used ENCODE transcription factor ChIP-seq data to identify HS risk variants at TF binding sites. We found 85 HS risk variants at binding sites for 198 TFs across 12 GWAS loci. Hierarchical clustering of TFs and loci revealed 17 TFs with binding sites across multiple GWAS loci, including TFs implicated in skin inflammation such as *STAT3*, *GATA3*, *CEBPB*, *NR3C1*, *FOS*, and *JUND* (**Figure 1C; Supplementary Data 3**). To assess whether any of these TFs are relevant to HS, we queried their expression in HS lesional and healthy skin using scRNAseq data (*18*). Of the 17 prioritized TFs, 16 were differentially expressed in HS skin (**Supplementary Data 4**). Six were upregulated in one or more cell types in HS relative to controls (*ATF3, CEBPB, GATA3, MAX, SMARCA4, STAT3*), and 10 were downregulated in all cell types relative to controls (**Supplementary Data 4).** *STAT3*, which has binding sites at 7 GWAS loci encompassing 13 HS risk variants, is a known master regulator of inflammation-driven repair response (*19*), and contributes to stemness and proliferation *via* PI3K/Akt signaling (*20*).

Taken together, this integrative analysis identified 67 HS risk variants with evidence of regulatory function, supporting a model in which cREs coordinate gene expression across GWAS loci in HS-relevant cell types. This data highlights, in particular, dysregulated interactions between the immune system and activated epithelial cells and opens multiple putative mechanisms for understanding HS pathophysiology.

### HS risk variants are linked to genes dysregulated in lesional skin

We next sought to identify genes regulated by HS risk variants using eQTL and chromatin accessibility data. Using GTEx eQTL data (*21*) we found that 17 HS risk variants across 10 loci are associated with the expression of 40 transcripts (coding and non-coding), 14 of which show skin-specific effects (**Figure 1D; Supplementary Data 5**; **Supplementary Figures 14-21**). Of these, four protein-coding genes - *INAVA*, *KIF21B*, *CYP1B1*, and *RBBP8* – showed colocalized signals between eQTL and HS GWAS loci, implicating them as direct targets HS risk variants. To improve detection power, we expanded our analysis to a larger blood-specific eQTL dataset (*22*), and identified 76 variants associated with expression of 40 transcripts, some of which were also identified in GTex (**Supplementary Data 6**). In total, across both eQTL datasets, 84 out of 214 HS risk alleles were annotated as eQTLs for 58 transcripts (**Table 3**). As a complementary approach, we used chromatin accessibility data from human terminal hair follicles (*23*) to investigate enhancer-gene interactions at HS loci. We identified 38 open chromatin regions (OCRs) containing one or more HS risk variants across 10 GWAS loci. These OCRs were statistically correlated with transcription start sites of 46 genes, implicating them as putative effector genes (**Supplementary Data 1 and 7**).

**Table 3.** eQTL and PheWAS Annotations of HS GWAS Risk Loci.

| Region | Transcripts carried on risk haplotype | Regulatory Targets | Drug Targets | Mendelian Disease Gene | GWAS catalogue variants | PheWAS |
| --- | --- | --- | --- | --- | --- | --- |
| **1q32.1** |  | CAMSAP2, DDX59, INAVA | CACNA1S |  | Celiac disease | **IBD and UC** |
| **2p22.2** | CYP1B1, CYP1B1-AS1 | ATL2, CYP1B1, RMDN2 | CYP1B1 |  | Nevus count, Nevus count or cutaneous melanoma | Skin cancer, Other nonepithelial skin cancer, Actinic keratosis, Degenerative skin conditions and other dermatoses |
| **2q22.1** | CXCR4, ENSG00000289974 | CXCR4 | CXCR4, DARS, LCT | CXCR4 | Interferon-related traits, monocyte count | - |
| **6p21.32<sup>†</sup>** |  | #N/A | CFB, C4B, CYP21A2, HLA-DRB1, PSMB8, PSMB9, COL11A2, RXRB | C4A, TAP1, TAP2, PSMB8, PSMB9 | rheumatoid arthritis, IgA nephropathy, asthma, type 2 diabetes, C-reactive protein levels, and IL-6 levels | IBD, UC, RA, T1D, CeD, MS, Chronic ulcer of leg or foot, Hypothyroidism, Polymyalgia Rheumatica |
| **9q31.3<sup>†</sup>** | ENSG00000231678 | - |  |  | - | - |
| **13q22.1-a** | KLF5 | MZT1, DIS3, PIBF1, KLF5 |  |  | Monocyte count | Varicose veins of lower extremity, varicose veins |
| **13q22.1-b<sup>†</sup>** | - | KLF12 |  |  | Allergic disease (asthma, hay fever or eczema), Asthma, Eczema, Hay fever and/or eczema, Pediatric asthma | Asthma, Diseases of esophagus, GERD |
| **14q24.3** | ENSG00000258740,<br>ENSG00000289340,<br>ENSG00000258820 | #N/A | NEK9,<br>PGF |  | - | - |
| **17q24.3<sup>†</sup>** | - | - |  | SOX9 | - | - |
| **18q11.2** | ENSG00000265943,<br>RBBP8, MIR4741,<br>RN7SL745P | CABLES1,<br>GATA6,<br>LAMA3,<br>RBBP8,<br>RIOK3,<br>TMEM241 | RIOK3,<br>NPC1,<br>LAMA3 |  | - | Pain |
| **18q12.2** | MAPRE2,<br>ENSG00000274400 |  |  |  | - | - |
| **22q12.1** | KREMEN1 | - | CHEK2 | KREMEN1 | - | - |
| **† previously associated with HS (Andersen et al., 2025)** |  |  |  |  |  |  |

Together, these analyses yielded 78 transcripts potentially regulated by HS risk variants (**Figure 1E**). By also including genes carried on risk haplotypes (*KREMEN1*) and implicated in previous HS GWAS (*WNT10A*, *NCSTN*, *PSENEN*), we selected 82 candidate genes for further investigation in HS skin. We queried single-cell RNA-seq data from HS lesional and healthy skin that profiles keratinocyte, fibroblast, melanocytes, smooth muscle, myeloid, mast, and T-cells (*18*). We identified 55 genes differentially expressed in at least one HS cell type (Table 3; **Supplementary Data 8**). Unsupervised hierarchical clustering of genes and cell types identified a strong gene signature in HS keratinocytes that includes 38 differentially expressed genes (DEGs), including upregulated *CXCR4*, *SOX9*, several *HLA* Class II genes, and *PSENEN*, among others (**Figure 1E; Supplementary Data 9**). Of the HS risk variants linked to keratinocyte genes, 36 are also located within candidate cREs (**Supplementary Data 10**), supporting the existence of a coherent transcriptional program that is disrupted by HS risk variants.

### CXCR4+ keratinocytes implicate MIF-mediated upregulated PI3K/AKT and NF-***κ***B signaling, which is pharmacologically validated

We selected *CXCR4* for further experimental studies in HS keratinocytes. First, it has been implicated in key features of HS pathology, including aberrant keratinocyte proliferation in response to inflammation (*24, 25*), pain perception (*26, 27*), and upregulated PI3K/Akt signaling Second, it also regulates stem and progenitor cell migration during development and oncogenesis, which is intriguing given the hypothesis that dermal tunnel formation in HS occurs via aberrant seeding of follicular stem cells in the dermis (*28*). Third, because multiple CXCR4 antagonists are either FDA-approved or in development for a variety of clinical immunology settings (*29*).

To characterize CXCR4’s role in HS pathogenesis we first performed immunohistochemistry and observed expression in epithelial tendrils and epithelialized dermal tunnels (**Figure 2A**), suggesting an association with aberrant keratinocyte migration. In healthy terminal hair follicles CXCR4 is expressed mainly in the epithelial outer root sheath (**Supplementary Figure 22**) and during development in epithelial cells of the hair follicle placode (*30*). We next examined the transcriptional profile of *CXCR4*+ HS keratinocytes by comparison to CXCR4-keratinocytes and discovered upregulation of GWAS genes including *SOX9*, *HLA-DQA*, *HLA-DQB1*, and *HLA-DRB* and dysregulation of TFs linked to HS risk variants (**Figure 2B**; **Supplementary Data 11**). Several other genes in the CXCR4+ keratinocyte gene signature have established roles in wound healing when keratinocytes acquire EMT characteristics to become migratory (*31*). Among these, *CD74* was upregulated and highly co-expressed with *CXCR4*. CD74 is a receptor for MIF that activates several pathways, including PI3K/Akt, ERK/MAPK, and NF-κB (*32*). Activation of these pathways is influenced by membrane localized protein-protein interactions. For example, heterodimerization of CD74 with CD44 induces MAPK/ERK signaling, while heterodimerization with CXCR4 amplifies PI3K/Akt signaling (*33*). CD74 activation of NF-κB occurs downstream of γ-secretase cleavage, which releases its intracellular domain (CD74-ICD) into the cytoplasm, where it interacts with RELA, translocates to the nucleus, and activates transcription of NF-κB target genes (*34, 35*). Co-expression analysis of *CXCR4*+ keratinocytes confirmed the presence of coreceptors (*CD44* and *CD74*), γ-secretase genes (*PSENEN* and *NCSTN*), and CD74-ICD binding partner, *RELA* (**Supplementary Figure 23**. Pathway analysis of genes differentially expressed in HS *CXCR4*+ keratinocytes compared to *CXCR4*-keratinocytes confirmed activated PI3K/Akt, ERK/MAPK, NF-κB, and P53 signaling in this cell type (**Figure 2C; Supplementary Data 11**), implying increased inflammation and suppressed apoptosis via CXCR4-CD74 signaling. Our data support a model in which HS keratinocytes are transcriptionally competent to exhibit aberrant activation of this inflammatory signaling axis.

**Figure 2.**
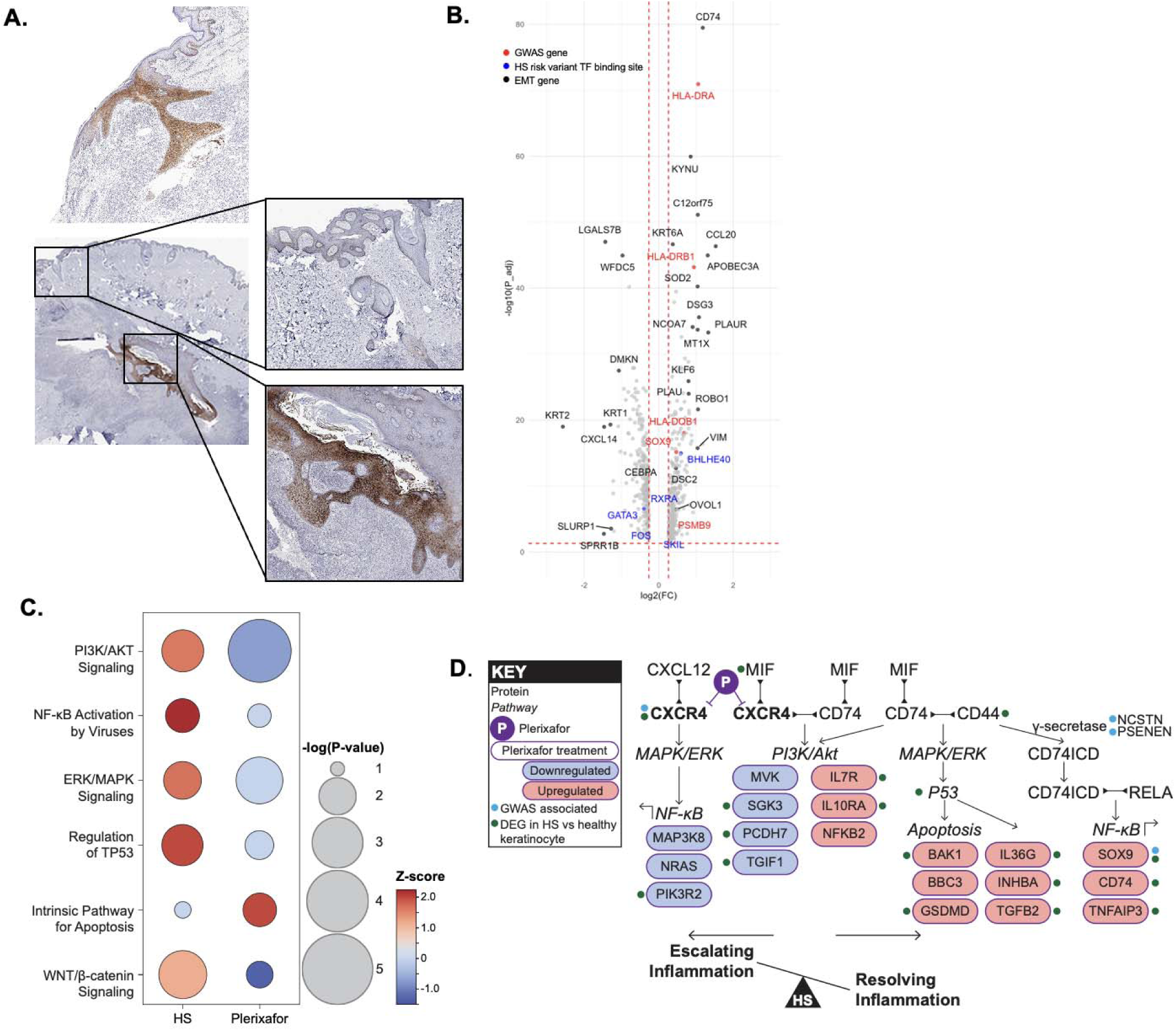
*CXCR4*+ keratinocytes in HS skin. (A). CXCR4 protein expression in HS skin. (B) We used single cell transcriptomic data to compare *CXCR4*+ keratinocytes to *CXCR4*-keratinocytes and found upregulated *SOX9* and HLA class II genes (red). We also identified transcription factors with binding sites that contain HS risk variants (blue) and a strong signature of epithelial mesenchymal transition associated with epithelial wound responses (black). Strong co-expression of CD74 suggest that CXCR4 may be forming a heterodimer with CD74 to amplify PI3K/AKT, ERK/MAPK, and NF-KB signaling. (C) Activation of these pathways is confirmed in CXCR4+ keratinocytes. Furthermore, exposure to Plerixafor in ex vivo HS skin attenuated these pathways in CXCR4+ keratinocytes. (D) Proposed model illustrating how upregulated *CXCR4* expression in HS keratinocytes disrupts the balance of inflammatory signaling pathways, and how Plerixafor treatment may restore this balance (purple). In this model, CXCR4 upregulation in HS keratinocytes promotes activation of MAPK/ERK, PI3K/Akt, and NF-κB pathways, contributing to sustained inflammation and impaired resolution. Pharmacologic inhibition of CXCR4 with Plerixafor attenuates these pathways and enhances apoptotic and gene programs associated with physiological remodeling in the hair follicle. Downstream pathway components that are modulated in CXCR4⁺ keratinocytes following Plerixafor treatment are highlighted in purple. Genes identified as GWAS-associated are marked with light blue indicators, and genes differentially expressed between HS and healthy keratinocytes are marked with green indicators.

CD74 plays a central role in mounting an appropriate inflammatory response, balancing activation of repair pathways without causing tissue damage (*32*). Thus, we hypothesized that in HS, upregulation of *CXCR4* in keratinocytes skews the balance towards sustained inflammation, excessive migration, and delayed repair programs (*33*). To test this, we performed ex vivo pharmacological inhibition of CXCR4 in HS skin with Plerixafor followed by single cell sequencing. Consistent with our hypothesis, CXCR4 inhibition (CXCR4i) attenuated PI3K/Akt and ERK/MAPK signaling, altered NF-κB pathway transcripts, and upregulated apoptosis pathways in CXCR4+ compared to CXCR4-HS keratinocytes (**Figure 2C**). CXCR4i also attenuated WNT/β-catenin signaling, a well-characterized marker of epithelial cell proliferation, and induced changes to genes previously implicated in NF-κB activation of epithelial downgrowth in physiological hair follicle remodeling in development and cycling that includes upregulated *SOX9*, *CD74*, and a central negative regulator of NF-κB mediated inflammation, *TNFAIP3* (**Supplementary Data 12**) (*36, 37*). These results support the dual role of NF-κB in epithelial tissues as both a driver of inflammation and a mediator of tissue repair (*38*).

Together our findings indicate that the MIF/CXCR4/CD74 axis is a key modulator of aberrant inflammation in HS keratinocytes, and that excessive *CXCR4* expression skews the inflammatory program towards pathological persistence rather than resolution. Our results also highlight CXCR4 as a potential therapeutic target in HS.

### Mendelian disease genes and drug targets at HS GWAS loci

GWAS loci are enriched for Mendelian disease genes that share clinical features and can provide insight into disease mechanisms and the effects of GWAS risk variants (*39*). Drug targets implicated by human genetic studies have also been shown to have higher drug development success rates (*40*). Accordingly, we examined whether any of our GWAS-linked genes are associated with any relevant Mendelian diseases or are known drug targets. Rare heterozygous gain of function variants in *CXCR4* cause WHIM (Warts, Hypogammaglobulinemia, Infections, Myelokathexis) syndrome, a single-gene disorder with variable expressivity that shares some clinical features with HS (*7, 41–44*). Beyond its pathogenic role in WHIM, *CXCR4* is implicated in other diseases and has therefore been pursued as a target for small molecule and peptide antagonists (*45*). Additional Mendelian disease genes that share some clinical features with HS are listed in Supplementary Information. Genes that are drug targets are listed in **Table 3** and described in **Supplementary Data 13**. Indications for GWAS-implicated drugs include HS comorbidities, such as hypertension, bacterial and viral infections, allergies, and irritable bowel syndrome.

### Disease associations implicated by shared polygenic architecture with HS

Polygenic architecture that is shared between diseases can contribute to the development of evidence-based screening recommendations for HS comorbidities, resolve disease mechanisms, identify drug repurposing opportunities, and improve drug safety profiles. To align the polygenic architecture of HS with that of other common diseases, we used variant-level and variant-aggregation methods. Specifically, we conducted PheWAS with individual HS risk variants and with an HS genome-wide polygenic risk score (GPS) that we developed and validated. We also conducted genetic correlation analysis using LD score regression.

### Variant Pleiotropy

We conducted a PheWAS analysis for each of the top variants associated with each locus in the eMERGE-III (*46, 47*), All of Us (AoU) (*48*), and UK Biobank datasets (*49*), followed by a meta-PheWAS across a total of 875,440 individuals (**Supplementary Figure 24**), which provided rigorous evidence for pleiotropy. This analysis confirmed several established HS comorbidities, including asthma, skin cancer, rheumatoid arthritis, and type 1 and 2 diabetes, among others. It also revealed new disease associations. For example, the HS risk allele rs41286076-C at the *KLF5* locus is associated with varicose veins. We also discovered a negative correlation between IBD and ulcerative colitis at chromosome 1q22.1 for a variant associated with expression of *INAVA* and *GPR25* (rs296539-C, OR=0.92, P=1.58E-06).

### Genome-wide polygenic risk score (GPS) development and PheWAS

We developed and evaluated an HS GPS using an independent dataset, UK Biobank (UKBB), consisting of European subjects (HS cases=213 and controls=381,611). This dataset was not included in the discovery of GWAS. The GPS demonstrating the best performance was established using the PRS-CSx method with phi=1.00E-04, encompassing variants across the entire genome (**Supplementary Data 14 and 15**).

Subsequently, we applied this GPS to subjects from the All of Us (AoU) release 2. Initially, we used it for Europeans within the AoU dataset (HS cases = 204 and controls = 35,326) (**Figure 3A**). The OR per standard deviation of GPS was 1.65 (95% CI: 1.44-1.90, P=8.35E-15). When comparing the top 1% vs the bottom 99% of the GPS, the OR was 12.8 (95% CI: 7.97-20.6, P=6.19E-26) (**Figure 4 (a) Supplementary Data 16**). Similarly, for research participants with African ancestry (cases=264, and controls=12,813), the OR per standard deviation of GPS was 1.26 (95% CI: 1.12-1.43, P=2.42E-04), and for the top 1%, the OR was 3.74 (95% CI: 1.87-7.51, P=1.98E-04). We also conducted tests on the Admixed American population (cases=72 and controls=9,827), yielding ORs of 1.41 (95% CI: 1.11-1.79, P=4.48E-03) and 1.49 (95% CI: 0.20-11, P=6.96E-01) per SD of GPS and for the top 1%, respectively.

**Figure 3:**
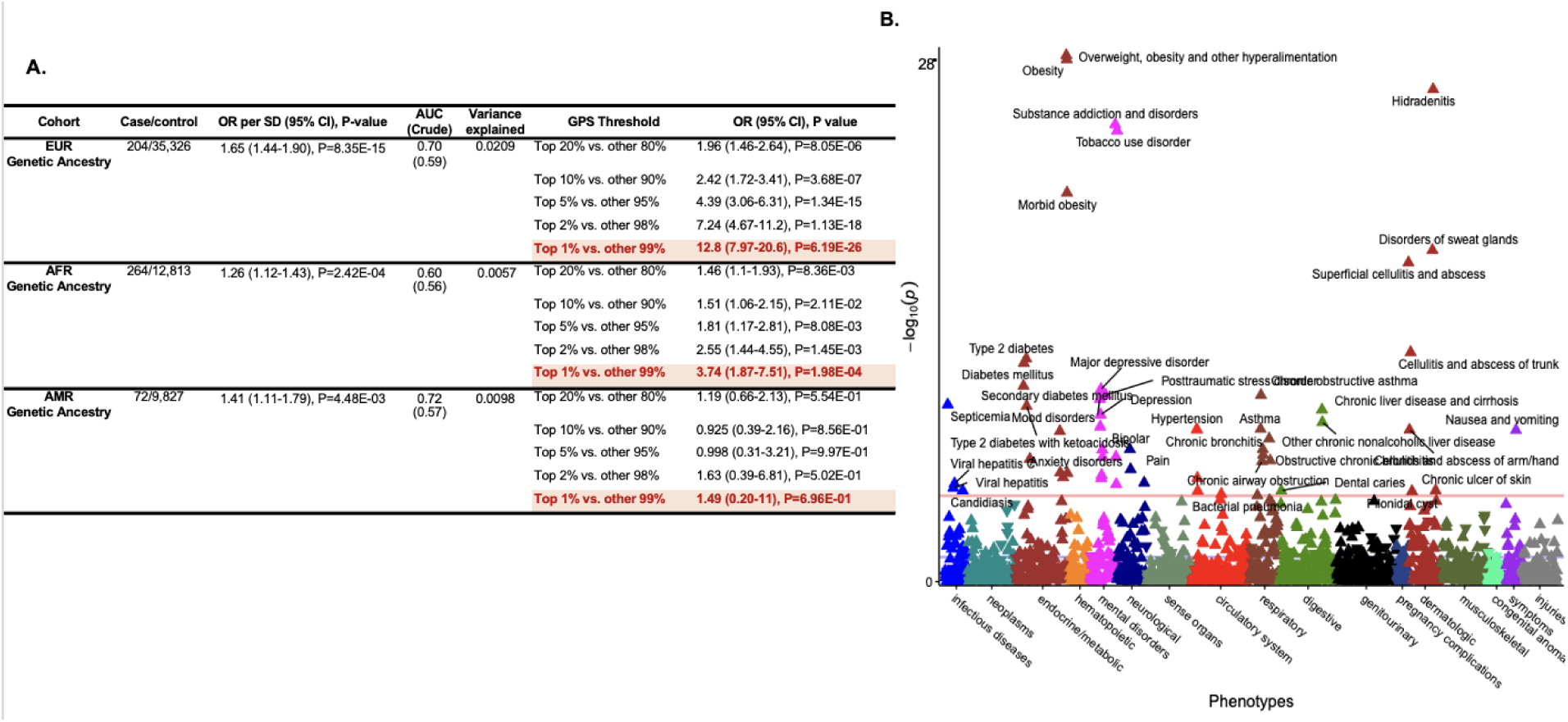
GPS testing results and PheWAS analysis in the AoU dataset. (a) **GPS testing results in the AoU Release 2 dataset.** The Odds Ratio (OR) corresponds to a model adjusted for age, sex, and principal components of ancestry. The Standard Deviation (SD) represents the GPS distribution in controls. The Area Under the Curve (AUC) indicates the performance of the model adjusted for age, sex, and principal components of ancestry. The “Crude” AUC refers to GPS performance without adjusting for covariates. The variance explained is calculated for the GPS component alone, without covariates. CI: Confidence Interval. (b) **PheWAS with HS GPS in the AoU dataset.** The analysis includes data from 148,234 AoU participants and was performed under a dominant inheritance model using logistic regression, adjusted for age, sex, and ancestry. Effect estimates and two-sided *P*-values were derived from logistic regression. The red horizontal line indicates the phenome-wide significance threshold after adjusting for multiple testing (P = 2.8 × 10⁻L). The Y-axis represents −log10(P-value), while the X-axis displays system-based phecode groupings. Upward-pointing triangles denote increased odds for a phecode, whereas downward-pointing triangles indicate reduced odds.

**Figure 4:**
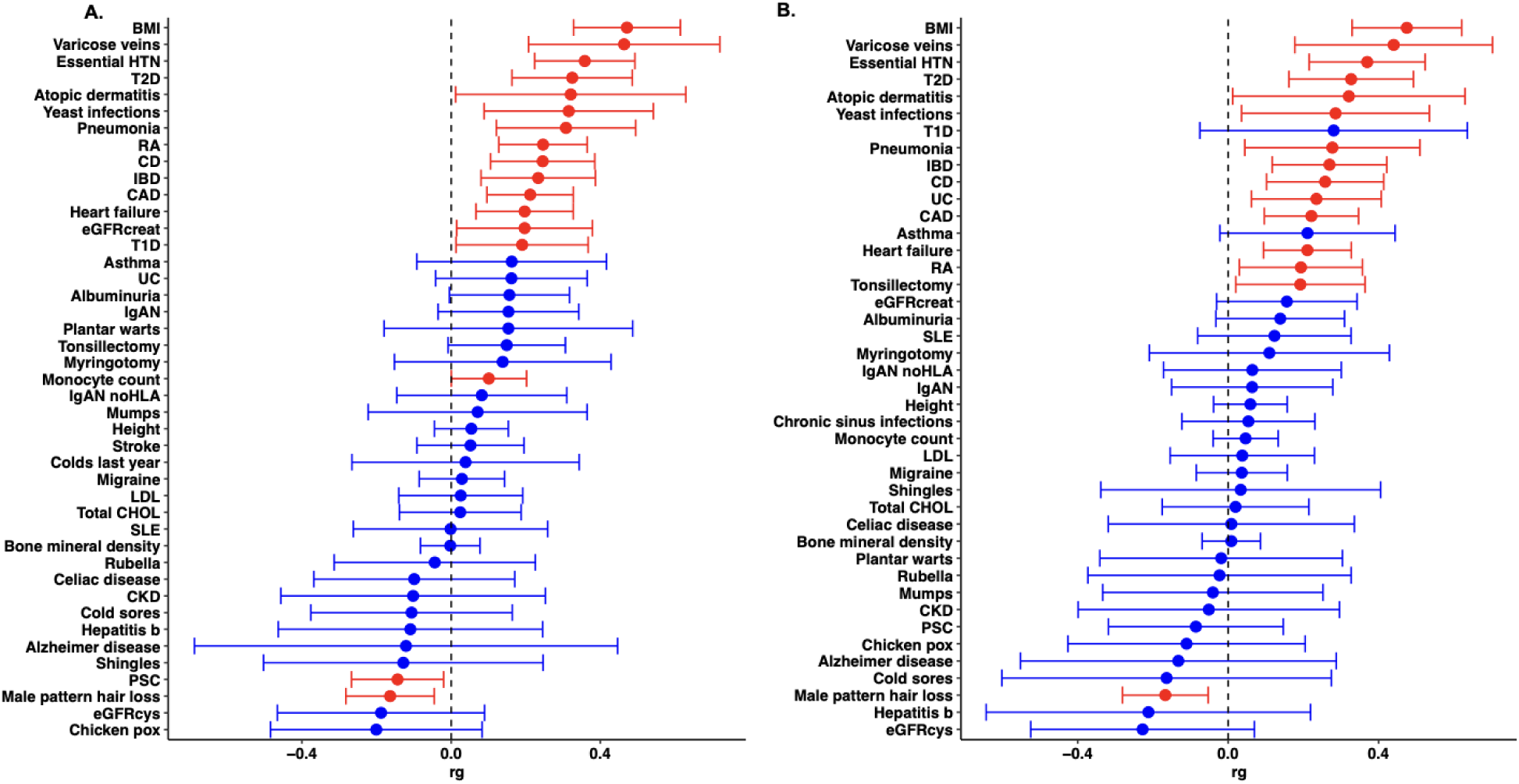
Genome-wide genetic correlations of GWAS results for HS with infectious, autoimmune, cardiometabolic, and hair disorders. (a) Genetic correlations including the HLA region. (b) Genetic correlations excluding the HLA region.

To further explore potential clinical effects of HS risk variants, we conducted a GPS-based PheWAS in the AoU dataset release 2, comprised of 148,234 unrelated individuals (**Figure 3B**). As expected, our GPS was positively associated with HS (OR=1.66, 95% CI:1.52-1.83, P=1.07E-26). We also identified previously established HS comorbidities such as obesity, type 2 diabetes, depression, asthma, and hypertension. We identified an association with Pain (PheWAS code 355.1), extending our single variant PheWAS results and suggesting a greater overlap in architecture. Several cardiovascular outcomes (e.g., myocardial infarction, atrial fibrillation) have important clinical implications, highlighting a need for proactive cardiovascular screening and risk factor management in patients with HS. Infection susceptibility was implicated by multiple infections and diseases, and supports inherited immune dysregulation as a contributing factor for some individuals with HS (*7*). There were negligible effects when we repeated the analysis with an HS GPS that excluded the *HLA* locus.

Finally, we repeated both analyses (with and without *HLA* variants), controlling for the presence of an HS ICD code to evaluate contributions from collider bias since HS is associated with our GPS and several diseases through epidemiological studies. Associations persisted with similar effect estimates (less than 10% difference) for most diseases. Interestingly, the adjusted model identified associations with inflammatory and toxic neuropathy (OR=1.10, p=5.5E10-6), and polyneuropathy in diabetes (OR=1.11, p=3.6E10-6), which were not associated with the HS GPS in the unadjusted model. This suggests that these neurological conditions may be underdiagnosed among individuals receiving clinical care for HS.

#### Polygenic Correlations

Finally, we conducted genome-wide genetic correlation (r_g_) analyses using LD score regression to examine shared susceptibility for paired traits (**Figure 4; Supplementary Data 17**). This analysis confirmed several PheWAS results, including the positive associations with varicose veins, infection susceptibility (e.g., yeast infections and pneumonia), and cardiovascular outcomes. A few r_g_ estimates were influenced by the HLA region. Monocyte counts, Type I Diabetes, and Primary Sclerosing Cholangitis are only significant when the HLA is included in the analysis, suggesting that r_g_ estimates are being driven by shared HLA risk alleles. The ulcerative colitis (UC) association was only significant when the *HLA* locus was excluded, suggesting that shared risk variants in the *HLA* have opposing effects in UC and HS. The *HLA* locus did not influence the association of either Crohn’s disease (CD) or IBD, which is consistent with minimal contributions from the *HLA* gene region to CD and IBD (*50*). Interestingly, we also identified a negative correlation with male pattern hair loss (MPHL), a trait that could not be investigated with PheWAS. The negative r_g_ is consistent with the recent *WNT10A* finding, where a single variant at this locus shows opposing effects in HS and MPHL (*10*), and further suggests more extensive sharing of opposing risk alleles.

#### Heritability Estimates

Due to sample size, SNP-based heritability could only be reliably estimated for individuals of European ancestry, and results indicated very low heritability (0.10%, 95% CI: 0.10%-0.20%). African American ancestry was similarly estimated to be low, but the wide CI limits confidence in this estimate (60%, 95% CI: 0.10-1.1%). The Admixed American subset (148 cases) was too small to estimate. Our estimates are much lower than estimates obtained with alternative methods (*51*). This discrepancy could be explained by considerable contributions from environmental risk factors or attenuation from disease misclassification that occurs when relying on phenotypes derived from electronic health records in biobanks (*52*).

## Discussion

We conducted the largest HS GWAS to date and identified 214 risk variants at twelve loci. We conducted further studies to begin defining the biological and clinical effects of these variants.

We identified gene regulatory programs active in multiple disease-relevant cell types that are likely contributing to a broader multicellular dysregulation. We performed experimental validation in keratinocytes. CXCR4, a co-receptor of CD74, emerged as a key genetic driver in HS pathogenesis. Our data support a model in which CD74 protein complexes in keratinocyte cell membranes influence epithelial cell state by regulating pathways that influence inflammation and cell proliferation (PI3K/Akt, WNT), and apoptosis (P53). Furthermore, NF-κB activity is calibrated to balance escalation and resolution of inflammation (**Figure 5**). On one hand, NF-κB signaling downstream of CXCR4 and CD74 promotes escalating inflammation by inducing inflammatory genes such as *MAP3K8*, *NRAS* and PIK3R2, as evidenced by their downregulation upon CXCR4 blockade. Conversely, NF-κB signaling through CD74 and CD44 – mediated by γ-secretase cleavage of CD74 – regulates genes important for physiological hair follicle remodeling and inflammation resolution, including *SOX9, CD74,* and *TNFAIP3*. These genes are upregulated as epithelial cells activate to initiate hair follicle downgrowth, both during early follicle development and then later during hair follicle cycling (*36, 37, 53, 54*). These genes are likewise induced upon CXCR4i in HS keratinocytes (**Figure 2D**). This invites further studies to determine if CXCR4-CD74 signaling could be an upstream activator of NF-kB signaling in the hair follicle, which remains poorly defined (*55*). Importantly, *TNFAIP3* is a potent inflammation inhibitor and a key component of a negative feedback loop in NF-κB signaling. The clinical relevance of *TNFAIP3* in HS pathogenesis is further highlighted by its role in causing HA20 haploinsufficiency, which sometimes co-presents with HS (*56*).

**Figure 5.**
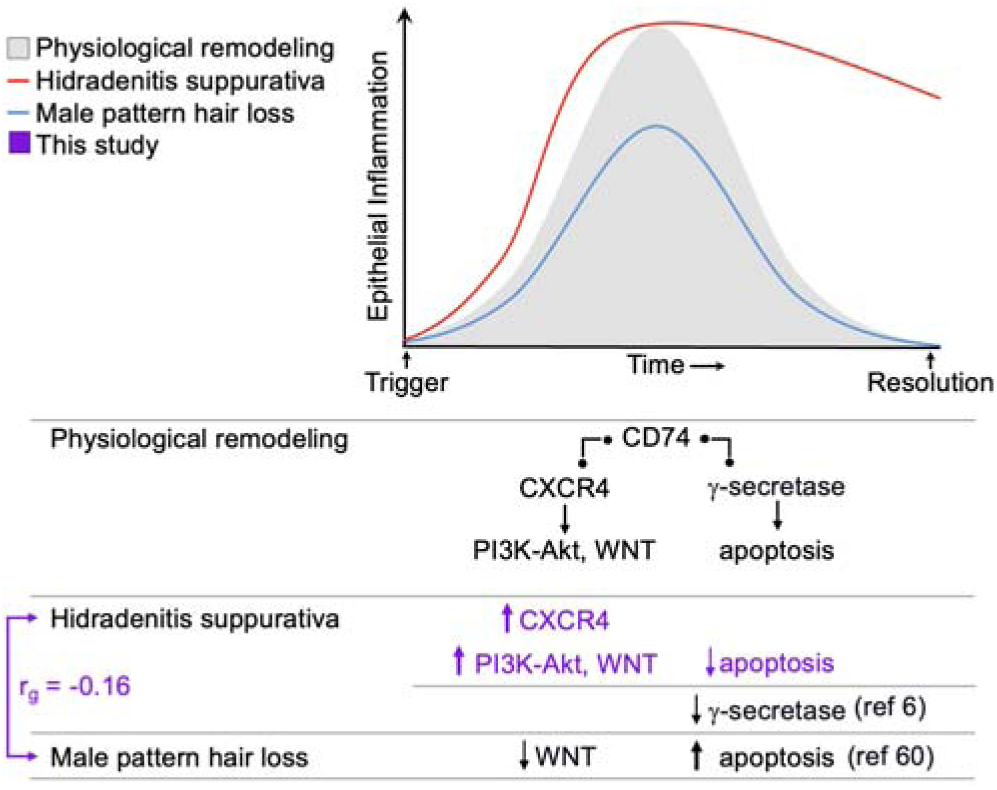
Genetic evidence supports CD74-mediated regulation of inflammation in HS and male pattern hair loss (MPHL). CD74 is critical for balancing inflammation to restore tissue homeostasis. Our genetic and experimental data support a model of aberrant sustained inflammation in HS keratinocytes driven by upregulated CXCR4 expression. CXCR4 blockade attenuated markers of aberrant inflammation and proliferation (PI3K/Akt, WNT signaling). γ-secretase processing of CD74 is upstream from genes that attenuate inflammation (e.g. *TNFAIP3*), instigate apoptosis (e.g., *BAK1*), and are implicated in epithelial activation and downgrowth in hair follicle development and cycling (*CD74, TNFAIP3, SOX9*). Our model suggests that loss of function variants in γ-secretase genes (*NCSTN, PSENEN*) would have the same effect as CXCR4 overexpression – escalating inflammation and preventing its resolution. The negative genetic correlation we identified between HS and MPHL further suggests that MPHL may result from an imbalance of this signaling axis in the opposite direction. Insufficient inflammation and increased apoptosis would result in attenuated epithelial downgrowth leading to hair follicle miniaturization.

Our data suggests that CXCR4 blockade restores homeostasis in keratinocytes and thus represents a new therapeutic strategy in HS. Importantly, CXCR4 blockade also disrupts CXCL12 signaling, targeting CXCR4’s chemotactic role and impairing immune cell trafficking in HS lesions. Thus, CXCR4 inhibition would have broad effects on restoring homeostasis in HS skin. While rare pathogenic variants in *CXCR4* were previously known to cause an Inborn Error of Immunity, our study expands human genetic evidence to also include common genetic variants that have more subtle phenotypic effects. This expanded evidence base increases our confidence in its therapeutic potential for HS management (*40, 57*). Our data also supports PI3K/Akt signaling inhibition as a viable therapeutic approach (**Figure 5**), which was first suggested by studies of *NCSTN* LOF and is also supported by the use of Sirolimus, an mTORC1 inhibitor, as an effective rescue therapy for HS (*7*).

Our model, which places CD74 at the fulcrum between inflammation escalation and resolution, also provides a new hypothesis for the effects of *NCSTN* and *PSENEN* loss of function (LOF) variants in HS. We propose that γ-secretase LOF impairs CD74 processing, which has the downstream effect of preventing inflammation resolution by perpetuating PI3K/AKT signaling (*58, 59*) and hindering *TNFAIP3* induction – conceptually mirroring the effects of *CXCR4* upregulation (**Figure 5**). Our model provides mechanistic coherency between our GWAS and familial genetic studies of HS.

We investigated the clinical effects of HS risk variants with several methods and discovered a genetic basis to comorbidities that have immediate clinical implications for HS patients, such as cardiovascular diseases and infection susceptibility. We also discovered a negative genetic correlation between MPHL and HS, which is consistent with the recent observation of a *WNT10A* low frequency missense variant that is inversely associated with both traits (*10*). We show that WNT signaling was activated and apoptosis suppressed in HS keratinocytes, and that CXCR4 inhibition reversed these effects. WNT signaling and apoptosis are also implicated in MPHL by GWAS (*60*), suggesting that MPHL may be characterized by imbalance of the same CXCR4–CD74 axis as in HS, albeit in the opposite direction. Insufficient proliferation and increased apoptosis would attenuate epithelial downgrowth thereby promoting hair follicle miniaturization (**Figure 5**). Supporting our hypothesis, upregulated PI3K/Akt signaling is emerging as a potent effector in promoting hair growth (*61–63*).

Limitations of our study include the use of biobank data. For example, the ICD-9 and -10 diagnosis codes for HS have very high sensitivity, but low specificity (*52*). This increases false positives among controls and may attenuate power to detect associations. Also, a lack of HS specific enhancer-target maps limits our ability to accurately map risk variants to transcriptomic changes. Likewise, eQTL effects may be missed in cell types that are not well-represented in publicly available data. Finally, further research into the polygenic architecture of HS is needed. Larger cohorts will allow us to identify additional risk variants, which in turn will allow us to define pathogenically relevant cell types with greater precision and to further improve the performance of a HS GPS.

Together our findings provide important insights into the genetic and cellular mechanisms linking HS risk variants to disease pathology, implicating the CXCR4-CD74 axis as a central regulator of pathological inflammation and highlighting novel therapeutic opportunities based on restoring tissue homeostasis in HS.

## Methods

### Meta-analysis

We conducted a meta-analysis by pooling data from 10 distinct trans-ancestry cohorts. Participants encompassed three major ancestry groups: European (4,100 cases and 1,311,317 controls), African American (2,113 cases and 185,901 controls), and Admixed American (148 cases and 19,611 controls). Cohorts include biobanks and a clinical cohort (Table 1). All participants provided informed consent to participate in genetic studies.

Our analytical approach involved initially analyzing each cohort independently. Subsequently, we combined the cohort-specific results using a fixed-effects meta-analysis method.

#### Electronic Medical Records and Genomics (eMERGE)

The eMERGE network provides access to electronic health record (EHR) information linked to GWAS data for 102,138 individuals recruited in three phases (eMERGE-I, II, and III) across 12 participating medical centers in years 2007-2019 (54% female, mean age 69 years, 76% European, 15% African American, 6% Latinx, 1% East or Southeast Asian by self-report). All individuals were genotyped genome-wide, and details on genotyping and quality control analyses have been described previously (*46, 47*)

#### All of Us (AoU)

The AoU research program launched recruitment in 2018 across 340 sites across the United States, and over 372,380 participants were enrolled by 2022. AoU combines participant-derived data from surveys such as self-reported health information, physical measurements, electronic health records, and biospecimens. We analyzed the AoU data on Workbench, a cloud-based environment(*48*). The first data release included N=98,622 participants with complete SNP microarray and genome sequencing data as well as phenotype information. The participants included 60% female, the mean age was 55 years and consisted of 53% European, 4% Asian, and 21% Black/African American race by self-report.

#### UK Biobank (UKBB)

UKBB is a prospective cohort based in the United Kingdom that enrolled individuals ages 40-69 across the UK in years 2006-2010(*49*). This cohort comprised of 488,377 individuals (54% female, mean age 57 years, 94% European, 2% East or Southeast Asian, and 2% African ancestry by self-report), genotyped with high-density SNP arrays, and linked to EHR data. All individuals underwent genotyping with UK Biobank Axiom array from Affymetrix and UK BiLEVE Axiom arrays (∼825,000 markers). Genotype imputation was carried out using a 1000 Genomes reference panel with IMPUTE4 software, as previously described(*49*). We then applied QC filters similar to eMERGE-III, retaining 9,233,643 common (MAF > 0.01) variants imputed with high confidence (R**_2_** > 0.8). For PCA by FlashPCA(*64*), we used a set of 35,226 variants that were common (MAF ≥ 0.01) and pruned using --indep-pairwise 500 50 0.05 command in PLINK v.1.9(*65*). *APOL1* G1 and G2 alleles were imputed separately using the TOPMed imputation server(*66*).

#### UK & Ireland HS GWAS Study

For participants recruited at Guy’s and St. Thomas Trust, ethical approval of human participant research was granted by the College Research Ethics Committee. For Dublin research participants, ethical approval was granted by the Health Product Regulatory Authority in Ireland. Written informed consent was obtained from all participants prior to participating in this study. Data were anonymized and handled in compliance with the UK General Data Protection Regulation (UK GDPR) and institutional policies governing the use of human genetic data. DNA samples were genotyped on Illumina Infinium Global Screening Array + Multi Disease (version: GSAMD-24v3-0-EA_20034606_A2), with 730,059 SNPs initially genotyped. Additional details are in Supplementary Information.

### Phenotyping of HS

Hidradenitis Suppurativa (HS) cases were defined using specific International Classification of Diseases (ICD) codes, including ICD-9 code 705.83 and ICD-10 code L73.2. Cases were identified by having one or more HS ICD codes, a method that has demonstrated high sensitivity (97-100%) and specificity (86-91%) (*67, 68*). Controls were identified by having no HS ICD codes in their records and were derived from the same populations as the cases. This phenotyping approach was applied across all cohorts to ensure accurate case identification in each cohort.

### Association and meta-analyses

Individual study cohorts include ethics, recruitment, genotyping, imputation, and quality control analyses, and the details of each separate cohort are described in **Supplemental Note**. We conducted genome-wide association analyses in the 10 cohorts using imputed dosage data under an additive model with adjustment for significant principal components (PCs) in each cohort with REGENIE or SAIGE software (*69, 70*). Only high-quality (R2 > 0.30) common (MAF > 0.01) single Nucleotide Polymorphisms (SNPs) were included in GWAS. Subsequently, we performed meta-analyses using the fixed effects inverse-variance-weighted method METAL(*71*). Genome-wide distributions of P-values were examined using quantile-quantile (QQ) plots for each cohort and the combined analysis. We only keep those SNPs typed or imputed in greater than or equal to 50% in cohorts. We defined the genome-wide significance threshold at P < 5.0E^-^ 08.

### Conditional analyses

To identify independent associations at specific loci, we carried out stepwise conditional analyses using the conditional and joint (COJO) association analysis method available in GCTA with an external 1000 Genome phase 3 reference panel(*72–74*). These conditional analyses were conducted with a significance threshold of P ≤ 5.0E-08 and using an LD reference based on data from the 1000 Genomes Phase 3, encompassing European and African American populations(*74*). We continued these conditional analyses for markers with a conditioned P ≤ 5.0E-^08^ until no remaining genome-wide significant associations were detected.

### Heritability and genetic correlations

We estimated SNP-based heritability using LD score regression (LDSC), using reference data from the combined 1000 Genomes phase 3 European and African American populations(*75*). We computed genetic correlations using bivariate LD score regression to explore potential shared genetic effects between HS and other traits. We collected summary statistics from the largest available GWAS for each phenotype. Summary statistics for immune and cardiometabolic traits were obtained from the LD Hub (*76*), GWAS catalog (*76*), and our previous GWASs (*77, 78*). Additionally, summary statistics for infection-related phenotypes were provided by 23andMe (*79*).

### Functional annotations of variants and loci

We used used the CARMA software to compute 95% credible sets for significant loci; these sets were expanded to incorporate further variants exhibiting strong linkage disequilibrium analysis (LD; r2 ≥ 0.8) with the lead SNP based on estimates from the 1000 Genomes Project. We used this list of SNPs to perform functional annotation as follows. Coding variants and their predicted functional consequences were identified using ANNOVAR. Regulatory activity of was assessed using the FUN-LDA model, following the approach in Gazal et al., 2018 (reference). To infer putative target genes variants were compared against expression quantitative trait loci (eQTL) datasets including: the Database of Immune Cell eQTLs (DICE) project, blood eQTLs derived from the eQTLGen consortium (*22*), tissue eQTLs and splicing quantitative trait loci (sQTLs) from the Genotype-Tissue Expression (GTEx) project (*80*). Differential expression analysis was performed to assess the expression of genes identified as being associated to GWAS variants across HS tissues compared to healthy controls using single cell RNA sequencing data from Gudjonsson et al. 2020 (GSE154773) and under accession number PRJNA1054546. COLOC software was used to perform colocalization analysis, with a threshold of PP4 > 0.5 considered indicative of shared causality (*81*). Transcription factor binding sites overlapping GWAS variants were identified using ENCODE3 ChIP data (retrieved via the UCSC Table Browser on July 10, 2024). Binding sites were filtered to include only TFs upregulated in one or more HS cell types, according to published scRNAseq data from HS tissues (*18*).

### Annotations from scATAC-seq and scRNA-seq in human skin with terminal hair

Cell Ranger outputs of 10 scalp scRNA and 9 scalp scATAC datasets were obtained from GEO accession ID GSE212450. Analysis was performed using R version 4.2.0. Quality control and downstream analysis of scRNA and scATAC, along with integration analysis, were conducted following the methodology previously described (*23*). HS GWAS risk variants were used to query open chromatin regions (OCR). The ‘addPeak2GeneLinks’ function in ArchR was employed to link genes to peaks(*82*), and high-confidence peak-gene links were obtained by filtering those with p2gCorr < 0.5. Subsequently, we identified HS risk variants overlapping with the peak regions. For OCRs containing HS risk variants but without correlated TSS, we identified plausible candidate genes under the assumption that those OCRs contain long-range enhancers for genes that are differentially expressed in the disease. The detection limit for the method we used to correlate OCR with TSS is 250Kb. Thus, we identified genes that are differentially expressed in HS lesional skin versus controls (GSE154773, PRJNA1054546)(*18*) and located between 250Kb and 800Kb distal to the OCR. We did not use this method to investigate long-range enhancers at chromosomes for locus 6p21.32, as the complex LD structure of the HLA complicates the interpretation of results.

### Drug Target Annotations

We identified drugs and bioactive molecules that target implicated genes using DrugBank version 5.1.11 data(*83*) and ChEMBL Release 33 data.(*84*) Using DrugBank data, we first mapped our implicated genes to DrugBank gene products and then gene products to drug targets. We then filtered on those drugs which target the identified drug targets. Using ChEMBL, we identified compounds with evidence of drug-like activity by mapping from our genes to ChEMBL compound sequences, compound sequences to compound targets, compound targets to assays, and assays to biological activity results. We then selected those results with a pChEMBL value greater than 6 and selected all implicated molecules. pChEMBL values provide a comparable activity value between studies, defined as −log10 (molar IC50, XC50, EC50, AC50, Ki, Kd, or Potency).(*84*) Minimum pChEMBL values between 5 and 6 are standard thresholds used to define active compounds.(*85*) We pulled additional molecules using ChEMBL mechanistic data by mapping from our genes to component sequences, component sequences to target components, target components to targets, targets to drug mechanisms, and finally, drug mechanisms to molecules.

For both ChEMBL and DrugBank, we limited our targets to those found in humans. We pulled indication and ATC 17 information from DrugBank for all available compounds and drug availability information from both ChEMBL and DrugBank. We categorized molecules as “available” according to ChEMBL if the availability type provided by ChEMBL is “prescription only” or “over-the-counter.” We categorized molecules as “ever available” according to DrugBank if the drug is categorized as “approved” or “nutraceutical.” We say “ever available” because DrugBank compounds that are withdrawn in one jurisdiction but approved in another retain the approved annotation. We additionally flagged drugs that were withdrawn in one or more jurisdictions according to either DrugBank or ChEMBL.(*83, 84*)

### PheWAS

We conducted a comprehensive meta-PheWAS analysis by combining data from the eMERGE-III, All of Us, and UKBB datasets for each independent genome-wide significant locus. The eMERGE-III dataset comprised EHR-linked GWAS data for 102,138 individuals and AoU 165,208, while the UKBB dataset included 488,377 individuals with a similar data linkage setup. To enable a harmonized meta-analysis, we standardized the phenotype data by converting all available ICD-10 codes to the ICD-9-CM system. This conversion process revealed 20,783 unique ICD-9 codes for eMERGE participants, 12,945 for AoU, and 10,221 unique ICD-9 codes for UKBB participants. Subsequently, we mapped these codes to 1,817 unique phecodes and performed logistic regression analyses. Our regression models accounted for various covariates, including age, sex, study site, imputation batch, and five principal components reflecting ancestry information.

Phenome-wide associations were performed using the PheWAS R package(*86*). The package uses two ICD-9 codes occurrences within a given phecode grouping to define a case and pre-defined “control” groups for each phecode. All 1,817 phecodes were tested using logistic regression with case-control status as the outcome and genotype, sex, age, batch, and five principal components of ancestry as predictors. We set the Bonferroni corrected statistical significance threshold for phenome-wide significance at 2.75E-05 (0.05/1,817 phecodes tested).

### Genome-wide polygenic risk score design and optimization

We used individuals of European ancestry of the UKBB (213 cases and 381,611 controls) to optimize the polygenic component of the GPS. Optimization was performed by selecting the best model PRS-csx(*87*). We used the summary statistics for 9,703,058, 17,299,793, and 11,914,123 for EUR, AFR, and AMR from our HS GWAS and 1000 Genomes Project phase 3 LD reference panel(*88*). We first computed five candidate GPS using the PRS-CSx(*89*) algorithm across the following phi range (fraction of causal variants): 1.00, 1E-02, 1E-04, 1E-06, 1E-08.

Each of the five scores obtained was subsequently evaluated for their ability to discriminate HS cases from controls in the UKBB optimization dataset. This evaluation took place after adjusting for age, sex, and the five principal components (PCs) of ancestry. The best performance score was identified based on the maximum area under the receiver operator curve (AUC) and the highest fraction of explained variance. Standard normalization procedures were applied to this score, subtracting the control mean and dividing by the control standard deviation.

## Acknowledgments

This research is based on data from the Million Veteran Program, Office of Research and Development, Veterans Health Administration. This publication does not represent the views of the Department of Veteran Affairs or the United States Government.

This study presents independent research supported by NIHR BioResource Centre Maudsley, NIHR Maudsley Biomedical Research Centre (BRC) at South London and Maudsley NHS Foundation Trust and Institute of Psychiatry, Psychology and Neuroscience (IoPPN), King’s College London.

The Estonian Biobank Research Team are: Andres Metspalu, Lili Milani, Tõnu Esko, Reedik Mägi, Mait Metspalu, Mari Nelis and Georgi Hudjashov. Computations were performed at the High Performance Computing Center, University of Tartu.

The Trøndelag Health Study (The HUNT Study) is a collaboration between HUNT Research Centre (Faculty of Medicine and Health Sciences, NTNU, Norwegian University of Science and Technology), Trøndelag County Council, Central Norway Regional Health Authority, and the Norwegian Institute of Public Health. The genotyping in HUNT was financed by the National Institutes of Health; University of Michigan; the Research Council of Norway; the Liaison Committee for Education, Research and Innovation in Central Norway; and the Joint Research Committee between St Olavs hospital and the Faculty of Medicine and Health Sciences, NTNU. The genetic investigations of the HUNT Study are a collaboration between researchers from the HUNT Center for Molecular and Clinical Epidemiology (formerly known as the K.G. Jebsen Center for Genetic Epidemiology as of August 1st 2023), NTNU, and the University of Michigan Medical School and the University of Michigan School of Public Health. We thank HUNT participants for donating their time, samples, and information to help others; clinicians and other employees at Nord-Trøndelag Hospital Trust for their support and for contributing to data collection.

## Funding

VA grant CX-002452 (LW) VA grant CX001897 (AMH)

Million Veteran Program MVP002 and MVP086

National Institute for Health and Care Research (NIHR) (XD-H)

Maudsley Charity grant ref. 980 (BioResource capital equipment funding)

Guy’s and St Thomas’s Charity grant STR130505 (BioResource capital equipment funding)

Estonian Research Council grant TK214 (EBRT)

National Institute for Health (NIH) NIAMS K01AR075111 (LP)

National Institute for Health (NIH) NIAMS R01AR080796 (LP)

## Author contributions

**Conceptualization:** AK, JG, JDM, LP

**Methodology:** AK, PAG, YL, JG

**Validation:** AK, JG

**Formal Analysis**: AK, PAG, YL, EPP, LW, AMH, TGD, MDR, AHS, HH, MM, ND, JB, MS, JS, XDH, SF, RC,CHC, SH, BK, MTL, KK, EBRT, LFT, MGH, JC, FM, PS, KLB, NT, ODP, AB, SR, SG, AO, LCR, CH, ML, ESC, CPL, GH, CW, KK, LCT, JEG, KRS, LP

**Investigation:** AK, LFT, ML, BMB, KV, LP

**Resources:** AK, AMH, VAMVP, MDR, HH, MS, LFT, ML, BMB, KH, NT, SR, AO, CH, CPL, KK, LCT, JEG, KRS, LP

**Data Curation:** AK, LFT, ML, BMB, KV, LP

**Writing – Original Draft:** AK, LP

**Writing – Review & Editing:** AK, PAG, LFT, ML, BMB, KH, KLB, ODP, AO, CH, LCT, JEG, KRS, JDM, LP

**Visualization:** AK, PAG, YL, LP

**Supervision:** AK, AMH, MDR, HH, MS, ML, BMB, KV, NT, SR, AO, CH, CPL, KK, LCT, JEG, KRS, LP

**Project Administration:** ML, BMB, KH, LP

**Funding Acquisition:** AMH, MDR, HH, MS, BMB, KV, NT, SR, AO, CH, CPL, KK, LCT, JEG, KRS, LP

## Data and materials availability

All data in this study are either publicly available or will be deposited in public repositories. The HS skin scRNAseq datasets used in this study are previously published and available under accession numbers GSE154773 and PRJNA1054546. The scRNAseq data generated in this study will be deposited in GEO prior to publication. All other data are available in the main text or supplementary materials.

## Supplementary Materials

Supplementary Text

Figs. S1 to S24

Tables S1 to S17

References (*91–113*)

