## Supplementary Information for "The polygenic architecture of hidradenitis suppurativa reveals signaling mechanisms that implicate epithelial remodeling"

**Supplementary Information**

COHORTS

Here, we have provided detailed descriptions of each cohort, including relevant information such as phenotyping of HS, quality control procedures, and association analysis methods.

### eMERGE-III

The eMERGE network has provided access to electronic health record information linked to GWAS data for 102,138 individuals recruited in three phases (eMERGE-I, II, and III) across 12 participating medical centers between 2007 and 2019. The cohort demographics consisted of 54% female individuals, with a mean age of 69. Self-reported ancestry distribution included 76% European, 15% African American, 6% Latinx, and 1% East or Southeast Asian. All individuals underwent genome-wide genotyping; details regarding genotyping and quality control procedures have been previously documented. In summary, all GWAS datasets were imputed using the multiethnic Haplotype Reference Consortium panel via the Michigan Imputation Server(*1*). This imputation was carried out in 81 batches. After imputation, we retained only markers with a Minor Allele Frequency (MAF) of at least 0.01 and an imputation quality (R2) of at least 0.8 in at least 75% of the batches. This resulted in a total of 7,529,684 variants being retained for subsequent for GWAS. For Principal Component Analysis (PCA), we conducted FlashPCA on a set of 48,509 common variants (MAF ≥ 0.01) that were also independent (determined through pruning in PLINK with the --indep-pairwise 500 50 0.05 command)(*2, 3*). The imputation of the G1 and G2 alleles of APOL1 was carried out separately using the Trans-Omics for Precision Medicine (TOPMed) imputation server. Our analytical pipeline involved a combination of VCFtools v.0.1.13 and PLINK v.1.9(*3, 4*). We used the REGENIE software to analyze each ancestral group's GWAS.

### All of US (AoU)

The All of Us research program commenced recruitment in 2018, enrolling participants from 340 sites across the United States, with over 372,380 individuals enrolled by 2022. AoU gathers participant-derived data through surveys, encompassing self-reported health information, physical measurements, electronic health records, and biospecimens. We analyzed AoU data within the Workbench, a cloud-based computational environment. The first data release from AoU included a cohort of N=165,208 participants, providing both SNP microarray data and phenotype information. Within this cohort, 60% were female, the mean age averaged 55 years, and self-reported racial backgrounds comprised 53% European, 4% Asian, and 21% Black/African American.

All participants underwent genotyping using the Illumina Global Diversity Array (GDA), which encompasses 1,904,679 Single Nucleotide Variants (SNVs) and 44,172 insertions/deletions (indels). Our initial step involved conducting genome-wide imputation analysis on the Workbench platform. Before imputation, we applied quality control measures, excluding variants with a Minor Allele Frequency (MAF) less than 0.005 (comprising 671,685 variants) or a genotype missingness rate exceeding 0.05 (totaling 41,526 variants). For 96% of the SNPs, we transformed genomic positions from human GRCh38 to hg19. Subsequently, we implemented the TopMed pre-imputation quality control (QC) pipeline to rectify allele designations and eliminate variants with poor mapping quality. Following these QC procedures, we retained 1,191,468 variants for the imputation process. To optimize computational resources and expedite processing, we divided the cohort of 165,208 subjects with microarray data into eight equal-sized batches and performed imputation independently for each batch. After pre-phasing with EAGLE v.2(*5*), we imputed missing genotypes using Minimac4(*6*) and 1KG phase 3v5(*7*) reference panel. After imputation, we retained only markers with a Minor Allele Frequency (MAF) of at least 0.01 and an imputation quality (R2). Similarly, we used the REGENIE software to analyze each ancestral group's GWAS in the All of Us workbench.

UK & Ireland HS GWAS

#### Cohort

#### 223 HS research participants from Guy’s and St. Thomas’ Trust (GSTT; N=112) and Dublin (N=111) were consented to participate in this study. For participants recruited at Guy’s and St. Thomas Trust, ethical approval of human participant research was granted by the College Research Ethics Committee. For Dublin research participants, ethical approval was granted by the Health Product Regulatory Authority in Ireland. Written informed consent was obtained from all participants prior to participating in this study. Data were anonymized and handled in compliance with the UK General Data Protection Regulation (UK GDPR) and institutional policies governing the use of human genetic data. DNA samples were genotyped on Illumina Infinium Global Screening Array + Multi Disease (version: GSAMD-24v3-0-EA_20034606_A2), with 730,059 SNPs initially genotyped.

#### Pre-imputation QC

After standard genotyping QC(*8*), including removal of SNPs/individuals with <1% call rates, removal of SNPs with <1% MAF, conversion to plus strand, removal of individuals with a mismatch between recorded sex and genetic sex, removal of up to 2nd-degree relations, removal of SNPs out of Hardy-Weinberg equilibrium (P < 7.5×10-8), removal of heterozygosity outlier individuals and removal of ancestry principal component outlier individuals (up to 20 PC), 503,598 genotyped common SNPs remained for 199 European ancestry HS samples (NDublin=106; NGSTT=93).

#### Merging with controls and QC

The HS dataset was merged with genotyped controls from the English Longitudinal Study of Aging (ELSA), consisting of unselected participants from England. ELSA individuals were genotyped using HumanOmni2-5-8-v1-0-D, and we selected SNPs (n= 248,662) where identical oligonucleotide probes were used for both HS and ELSA genotyping. After merging datasets removing ELSA ancestry outliers (Fig. 1) and removing 5 ambiguous sex individuals from the ELSA dataset, 7,072 ELSA controls remained, bringing the total number of individuals in the case-control dataset to 7,271.

#### Imputation

Genotype imputation was performed using the Michigan server (panel: HRC 1.1), generating genotype dosages for 9,979,264 variants with MAF>0.1% and INFO>0.7.

#### Association testing

Genome-wide association testing was performed using [REGENIE/logistic regression in PLINK], using 3 ancestry PCs and sex as covariates (Fig. 2 and 3, delete as appropriate). Summary statistics were filtered to only include SNPs with MAF>1% and INFO>0.8 (n=6,680,604).

### Estonian Biobank (EstBB)

#### Cohort Description.

The Estonian Biobank (EstBB) is a population-based biobank with more than 200,000 participants (https://genomics.ut.ee/en/content/estonian-biobank). The EstBB project is being conducted according to the Estonian Gene Research Act, and all participants have signed a broad informed consent form(*9*).

Upon recruitment, the biobank participants completed a thorough questionnaire covering lifestyle, diet and clinical diagnoses defined according to the ICD10 coding. In addition, the biobank’s health database is regularly updated by linking with national electronic health registries (including hospital databases). The EstBB samples were genotyped at the Core Genotyping Lab of the Institute of Genomics, University of Tartu, using Illumina global screening arrays v1.0, v2.0 and v2.0_EST. Individuals whose sex defined based on X chromosome heterozygosity did not match the sex recorded in phenotype data were excluded from the analysis. Before imputation, variants were filtered by call rate < 95%, Hardy–Weinberg equilibrium *P* < 1 × 10^-4^ (autosomal variants only), and minor allele frequency < 1%. Paraphrasing was performed using Eagle v2.3 software, and imputation was performed using Beagle v.28Sep18.793 using an Estonian population–specific imputation reference panel built from 2,297 whole-genome sequencing samples(*10*).

#### GWAS

Hidradenitsis suppurativa cases were defined by the ICD10 code L73.2 and controls as having no HS diagnosed. All individuals are of European descent. Association tests for HS were performed using SAIGE v0.43.1 software for imputed variants with adjustment for age, sex, and first 10 PCs.

### HUNT Study

The Trøndelag Health Study (HUNT) is a population-based cohort study carried out at four time points over approximately 40 years (HUNT1 [1984-1986], HUNT2 [1995-1997] and HUNT3 [2006-2008] and HUNT4 [2017-2019]). All inhabitants aged 20 years and over residing in Trøndelag County in Norway were invited to participate. The surveys included clinical measurements, blood sampling, and questionnaires on general health measures and a broad range of self-reported diseases and symptoms. Participants were also linked to regional- and national health registries through a unique national identification number, which enabled hidradenitis suppurativa (HS) cases to be identified by searching for HS ICD codes.

Participants from HUNT2-4 were genotyped using one of four different Illumina HumanCoreExome arrays (HumanCoreExome12 v1.0, HumanCoreExome12 v1.1, UM HUNT Biobank v1.0 and UM HUNT Biobank v2.0). Genotype calling was performed with GenTrain v.2.0 in GenomeStudio v.2011.1 (Illumina). Samples with <99% genotype calls, with large chromosomal copy number variants, contamination >2.5% as estimated with BAF Regress (4), with genotypic and phenotypic sex discordance, and not of European ancestry were excluded, leaving 88,600 genotyped subjects. Genetic variants out of Hardy-Weinberg equilibrium (p-value <0.0001) were excluded.

Imputation was performed on samples of recent European ancestry using Minimac4 (v1.0.2, https://genome.sph.umich.edu/wiki/Minimac4) from the Haplotype Reference Consortium (HRC) panel (v1.1), resulting in 24.9 million SNPs (INFO>0.3).

The ICD9 code 705.83 and the ICD10 code L73.2 were used to define HS cases. There were 365 individuals in HUNT with at least one of the specified codes, with the remaining 86 486 individuals defined as controls. GWAS was run in SAIGE v1.0.3 (5), using sex, age, genotyping batch and 10 ancestry principal components as covariates. Variants with MAF >1.7e-05 were included in the analyses, and dosages were used for imputed variants.

#### Ethics

Participation in HUNT is based on informed consent, and the study has been approved by the Norwegian Data Protection Authority and the Regional Committee for Medical and Health Research Ethics in Central Norway (REK Reference number 2015/586).

**Acknowledgements**

The Trøndelag Health Study (The HUNT Study) is a collaboration between HUNT Research Centre (Faculty of Medicine and Health Sciences, NTNU, Norwegian University of Science and Technology), Trøndelag County Council, Central Norway Regional Health Authority, and the Norwegian Institute of Public Health. The National Institutes of Health financed the genotyping in HUNT; the University of Michigan, the Research Council of Norway, the Liaison Committee for Education, Research and Innovation in Central Norway, and the Joint Research Committee between St Olavs Hospital and the Faculty of Medicine and Health Sciences, NTNU. The genetic investigations of the HUNT Study are a collaboration between researchers from the HUNT Center for Molecular and Clinical Epidemiology (formerly known as the K.G. Jebsen Center for Genetic Epidemiology as of August 1^st^, 2023), NTNU, and the University of Michigan Medical School and the University of Michigan School of Public Health. We thank HUNT participants for donating their time, samples, and information to help others, clinicians, and other employees at Nord-Trøndelag Hospital Trust for their support and for contributing to data collection.

### VA Million Veterans Program

**Core Acknowledgements**

**MVP Program Office**

- Sumitra Muralidhar, Ph.D., Program Director

US Department of Veterans Affairs, 810 Vermont Avenue NW, Washington, DC 20420

- Jennifer Moser, Ph.D., Associate Director, Scientific Programs

US Department of Veterans Affairs, 810 Vermont Avenue NW, Washington, DC 20420

- Jennifer E. Deen, B.S., Associate Director, Cohort & Public Relations

US Department of Veterans Affairs, 810 Vermont Avenue NW, Washington, DC 20420

**MVP Executive Committee**

- Co-Chair: Philip S. Tsao, Ph.D.

VA Palo Alto Health Care System, 3801 Miranda Avenue, Palo Alto, CA 94304

- Co-Chair: Sumitra Muralidhar, Ph.D.

US Department of Veterans Affairs, 810 Vermont Avenue NW, Washington, DC 20420

- J. Michael Gaziano, M.D., M.P.H.

VA Boston Healthcare System, 150 S. Huntington Avenue, Boston, MA 02130

- Elizabeth Hauser, Ph.D.

Durham VA Medical Center, 508 Fulton Street, Durham, NC 27705

- Amy Kilbourne, Ph.D., M.P.H.

VA HSR&D, 2215 Fuller Road, Ann Arbor, MI 48105

- Michael Matheny, M.D., M.S., M.P.H.

VA Tennessee Valley Healthcare System, 1310 24th Ave. South, Nashville, TN 37212

- Dave Oslin, M.D.

Philadelphia VA Medical Center, 3900 Woodland Avenue, Philadelphia, PA 19104

- Deepak Voora, MD

Durham VA Medical Center, 508 Fulton Street, Durham, NC 27705

**MVP Co-Principal Investigators**

- J. Michael Gaziano, M.D., M.P.H.

VA Boston Healthcare System, 150 S. Huntington Avenue, Boston, MA 02130

- Philip S. Tsao, Ph.D.

VA Palo Alto Health Care System, 3801 Miranda Avenue, Palo Alto, CA 94304

**MVP Core Operations**

- Jessica V. Brewer, M.P.H., Director, MVP Cohort Operations

VA Boston Healthcare System, 150 S. Huntington Avenue, Boston, MA 02130

- Mary T. Brophy M.D., M.P.H., Director, VA Central Biorepository

VA Boston Healthcare System, 150 S. Huntington Avenue, Boston, MA 02130

- Kelly Cho, M.P.H, Ph.D., Director, MVP Phenomics

MVP Core Acknowledgements for Publications_June 2025VA Boston Healthcare System, 150 S. Huntington Avenue, Boston, MA 02130

- Lori Churby, B.S., Director, MVP Regulatory Affairs

VA Palo Alto Health Care System, 3801 Miranda Avenue, Palo Alto, CA 94304

- Scott L. DuVall, Ph.D., Director, VA Informatics and Computing Infrastructure (VINCI)

VA Salt Lake City Health Care System, 500 Foothill Drive, Salt Lake City, UT 84148

- Saiju Pyarajan Ph.D., Director, Data and Computational Sciences

VA Boston Healthcare System, 150 S. Huntington Avenue, Boston, MA 02130

- Robert Ringer, Pharm.D., Director, VA Albuquerque Central Biorepository

New Mexico VA Health Care System, 1501 San Pedro Drive SE, Albuquerque, NM 87108

- Luis E. Selva, Ph.D., Director, MVP Biorepository Coordination

VA Boston Healthcare System, 150 S. Huntington Avenue, Boston, MA 02130

- Shahpoor (Alex) Shayan, M.S., Director, MVP PRE Informatics

VA Boston Healthcare System, 150 S. Huntington Avenue, Boston, MA 02130

- Brady Stephens, M.S., Principal Investigator, MVP Information Center

Canandaigua VA Medical Center, 400 Fort Hill Avenue, Canandaigua, NY 14424

- Stacey B. Whitbourne, Ph.D., Director, MVP Cohort Development and Management

VA Boston Healthcare System, 150 S. Huntington Avenue, Boston, MA 02130

Quality controls (QC), Imputation, GWAS and Meta-analysis

**Quality Control (QC) Filters Before Imputation:** We applied the following QC filters before proceeding with imputation:

**SNP QC:**

- Minor Allele Frequency (MAF) ≥ 0.01
- Genotype missingness (geno) ≤ 0.05 (Plink Threshold: <https://zzz.bwh.harvard.edu/plink/thresh.shtml>)
- Hardy-Weinberg Equilibrium (HWE) p-value > 1E-08 (for controls only)

**Sample QC:**

- Individual missingness (mind) ≤ 0.10
- Kinship analysis to detect and remove duplicate samples

**Imputation Process:** We utilized the Michigan Imputation Server and followed the data preparation guidelines provided (https://imputationserver.readthedocs.io/en/latest/prepare-your-data/) for imputation if not imputed the cohort(*11*).

**Population Stratification:** To account for population stratification, we divided the cohort into distinct ancestral groups (e.g., European, Hispanic, African, Asian) using global Principal Component Analysis (PCA). Precise genetic ancestry definitions were generated using machine learning algorithms like Random Forest.

**Post-Imputation QC Filters (for each ancestral group):** After imputation, we applied the following QC filters for each ancestral group:

- SNP QC: MAF ≥ 0.01, imputation R^2 ≥ 0.80

**PCA Analysis:** PCA analysis was performed after applying the above QC filters to generate ancestral group-specific principal components (PCs).

**GWAS Analysis:** Our GWAS analysis was conducted using the REGENIE package, which can accommodate related individuals. We separately applied the standard association model to each major ancestral group:

Y (Phenotype) = SNP dosage + Age + Sex + First 3 significant ancestry PCs (or any significant PCs).

These analyses were performed separately for each major ancestral group (e.g., European, Hispanic, or African).

**Meta-analysis:** In the meta-analysis phase, we conducted QC checks on each summary statistic received for every cohort. To ensure consistency across cohorts, we converted all data from hg38 to hg19 and verified that the tested MAF was consistent. Meta-analysis was performed separately for each major ancestry group and each dataset using a fixed-effect model with the METAL software(*12*).

### **Mendelian Disease Genes at HS GWAS loci**

GWAS loci are enriched for Mendelian disease genes that share phenotypic overlap (*13*) and can help refine knowledge about disease-relevant clinical features and interpret the effects of GWAS risk variants. Several genes prioritized by our GWAS are causes of Mendelian diseases that cause immune dysregulation (*C4A*, *TAP1*, *TAP2*, *PSMB8*, *PSMB9*, and *CXCR4*) (*14*). All six of these genes are upregulated in HS lesional skin in multiple cell types. Complete C4 deficiency is associated with increased susceptibility to infections with encapsulated organisms and lupus like disease (*15*). There are other monogenic disorders of the complement pathway which result in increased complement production/deposition. Defects in complement signaling have been implicated in HS (*16*), and there is evidence that this pathway may only be etiologically relevant for a subset of HS patients (*17*). MHC Type I deficiency is caused by autosomal recessive variants in *TAP1* and *TAP2* and is marked by an elevated risk of pyoderma gangrenosum, cutaneous granulomas, and sinopulmonary infections. Loss of function (LOF) variants in *PSMB8* or *PSMB9* cause CANDLE (chronic atypical neutrophilic dermatitis with lipodystrophy) syndrome, a recessive autoinflammatory interferonopathy that leads to a gain of NF-κB and type I interferon responses. Cutaneous features include neutrophilic dermatosis and sparse hair. WHIM (Warts, Hypogammaglobulinemia, Infections, Myelokathexis) syndrome is caused by heterozygous GOF variants in CXCR4 and is notable for highly variable penetrance and expressivity. Clinical hallmarks include significant immune dysregulation, even in the absence of infection, recurrent bacterial infections, susceptibility to viral infections, neutropenia due to impaired neutrophil egress from the bone marrow, low B cell number, and hypogammaglobulinemia (*18, 19*). Milder manifestations may be limited to autoimmunity (*20*). Cutaneous abscesses are reported (*21, 22*). It is interesting to note that pathogenic variants in *CXCR4* and *PSMB8* are associated with aberrant pigmentation, given the overlap of pigmentary disorders with familial HS attributed to g-secretase LOF (*14*).

Long-range enhancers located upstream from *SOX9* have been implicated in sex development disorders that affect gonad and/or genitourinary tract development and the endocrine-reproductive system (*23*). *SOX9* is a crucial regulator of hair follicle stem cell super-enhancers (*24*). *SOX9* has also been implicated in androgen signaling within the context of several cancers and by the identification of androgen receptor (AR) responsive enhancers at the *SOX9* locus. None of these defined long-range enhancers overlap with the LD block associated with HS. Thus, long range enhancer mapping within the context of HS is needed.

Ectodermal Dysplasia 13 (ECTD13) is a Mendelian disease caused by pathogenic variants in *KREMEN1*, located on the HS risk haplotype at chromosome 22q12.2. Ectodermal dysplasias are a heterogeneous group of hereditary diseases affecting ectodermal structures: teeth, hair, nails and sweat glands (*25*). Pathogenic *KREMEN1* variants cause alterations to WNT10A signaling in the Wnt/β-catenin signaling pathway (*26*). Patients experience sparse hair, abnormal scalp hair distribution, thin eyebrows and eyelashes, dry skin, low hairline, hyperhidrosis and heat intolerance (*26*). These phenotypes are similar to phenotypes seen in patients with *WNT10A* pathogenic variants (*27*).

### **Drug targets at HS GWAS loci**

Drug targets implicated by human genetic studies have greater drug development success rates (*28*). Identifying drug targets linked to GWAS risk variants may help to prioritize drug repurposing efforts and improve drug safety profiles (*28, 29*). Ten genes prioritized by functional annotations are identified as drug targets in DrugBank version 5.1.10 data^1^ and ChEMBL Release 20 data, including *CACNA1S*, *CXCR4*, *CYP1B1*, *DARS*, *HLA-DRB1*, *KLF5*, *LAMA3*, *PSMB8*, *PSMB9*, *RIOK3* (**Supplementary Data 13**). These genes are reported as targets for 48 different drugs, including immunomodulating agents (e.g., Fostamatinib, Nintedanib, Fedratinib, etc.), immunostimulants (Glatiramer, Plerixafor), anti-infectives (Zalcitabine, Framycetin), and calcium channel blockers (e.g., Diltiazem, Verapamil, etc). Indications for these drugs include hypertension, bacterial and viral infections, allergies, and irritable bowel syndrome, among others. Several of these indications are HS comorbidities (*14*).

**SUPPLEMENTARY FIGURES**

**Supplementary Figure 1:** Gene annotations and p-values for each independent genome-wide significant locus.

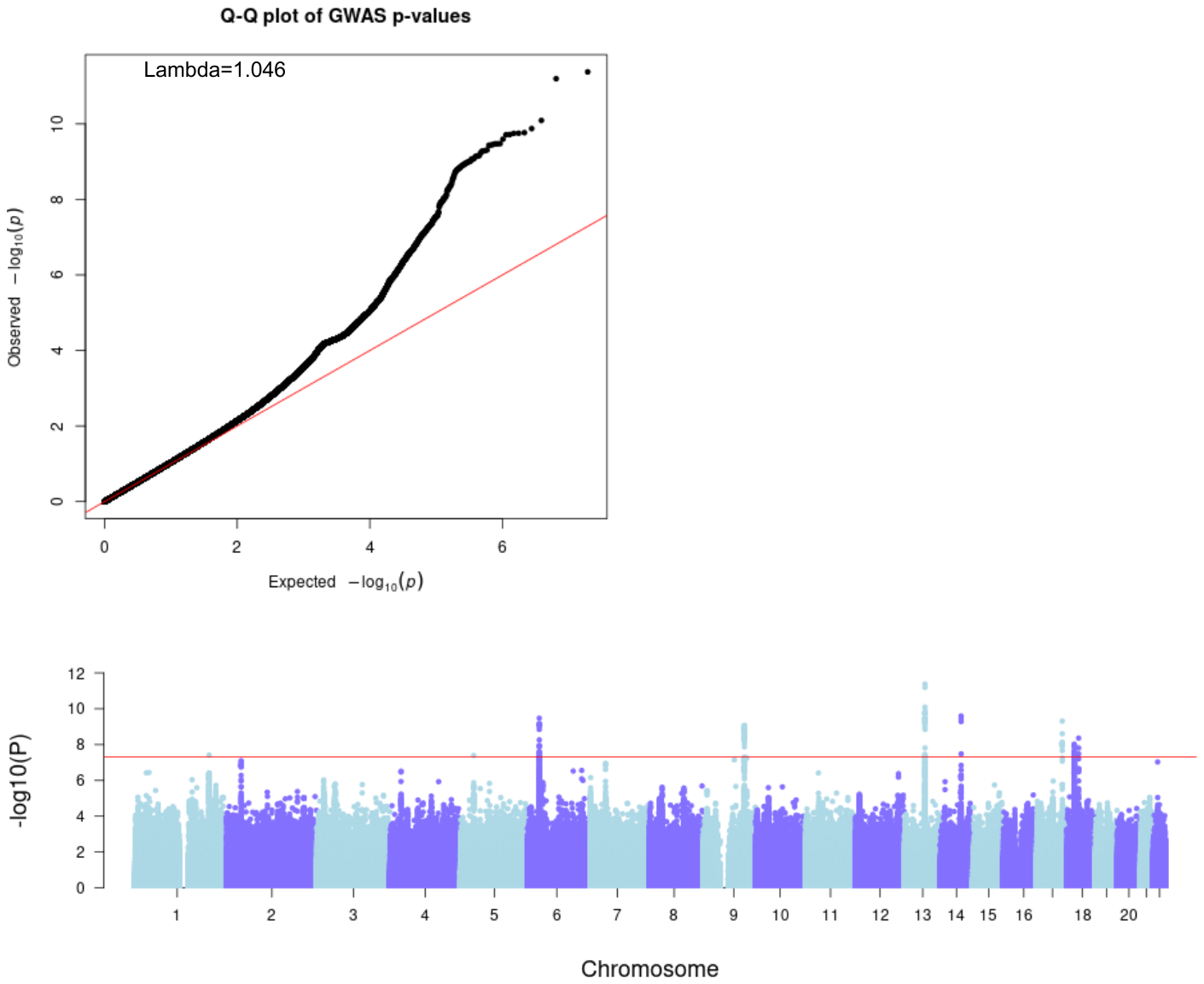

**Supplementary Figure 2:** Manhattan and Q-Q plots for the HS GWAS for European ancestry. The y-axis represents -log10 transformed P-values on both plots, while the x-axis represents genomic positions.

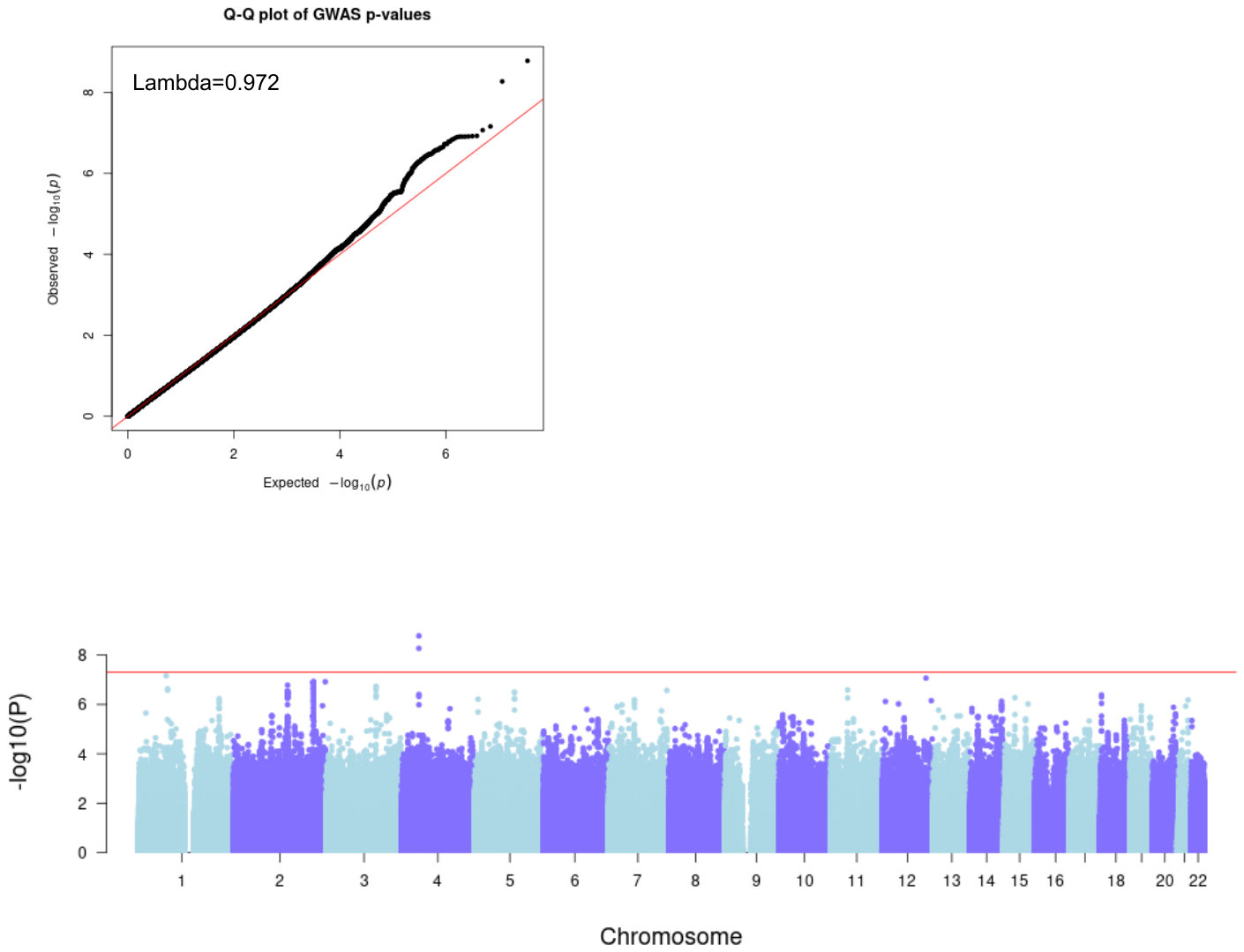

### **Supplementary Figure 3:** Manhattan and Q-Q plots for the HS GWAS of African Ancestry. The y-axis represents -log10 transformed P-values on both plots, while the x-axis represents genomic positions.

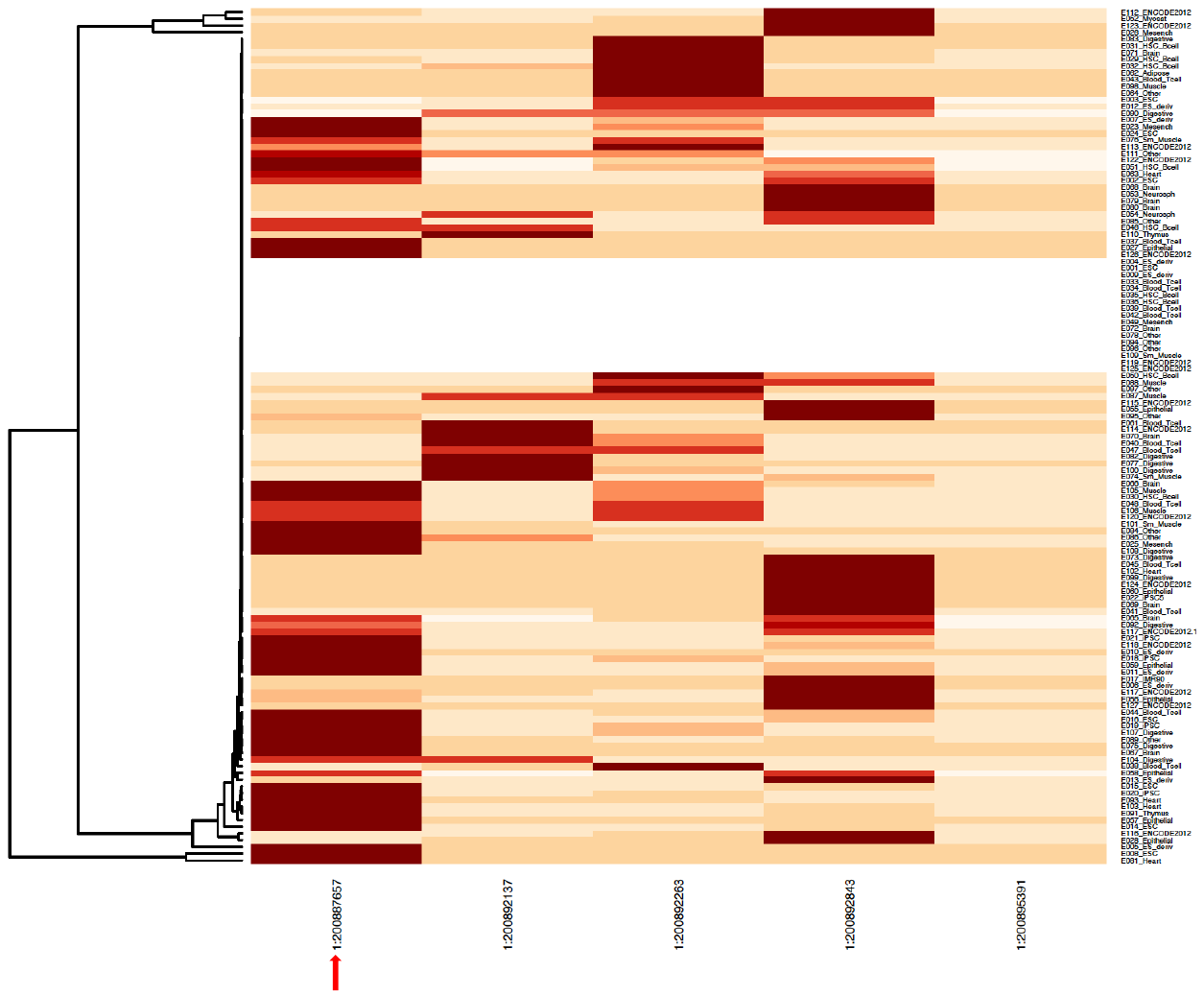

**Supplementary Figure 4:** Tissue-specific functional predictions were conducted for variants. The arrow indicates the index SNP (1:200887657). Additional columns along the X-axis represent all SNP proxies for the index variant, defined by having an R^2^ value of at least 0.8. These SNPs were arranged based on their genomic positions. The 127 tissues and cell types from the Roadmap Epigenomics project were clustered along the Y-axis using a hierarchical clustering approach. This clustering was based on the pattern of their functional latent Dirichlet allocation (FUN-LDA) scores. Darker colors indicate higher FUN-LDA scores, highlighting the functional significance of these variants in different tissues and cell types.

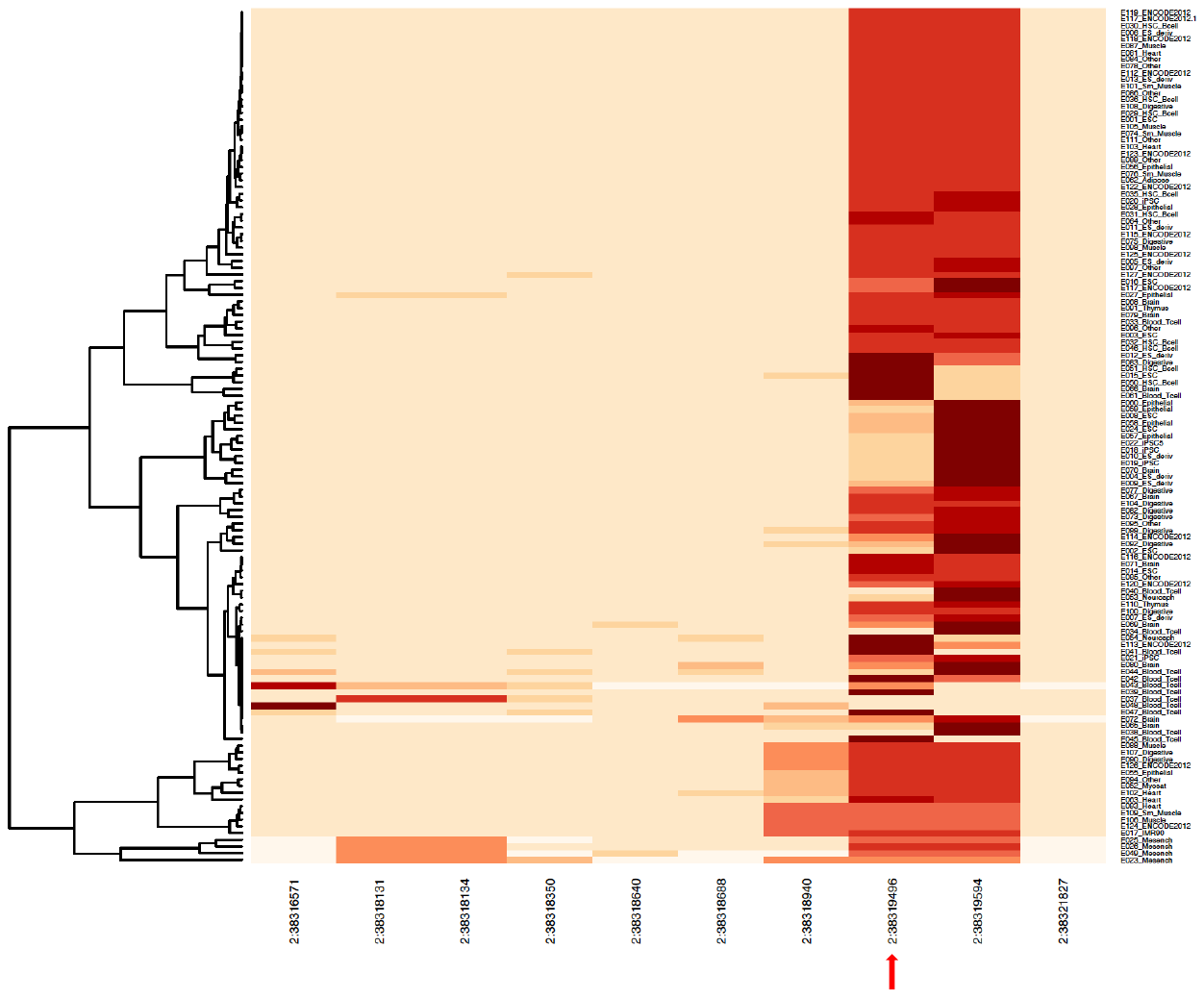

**Supplementary Figure 5:** Tissue-specific functional predictions were conducted for variants. The arrow indicates the index SNP (2:38319496). Additional columns along the X-axis represent all SNP proxies for the index variant, defined by having an R^2^ value of at least 0.8. These SNPs were arranged based on their genomic positions. The 127 tissues and cell types from the Roadmap Epigenomics project were clustered along the Y-axis using a hierarchical clustering approach. This clustering was based on the pattern of their functional latent Dirichlet allocation (FUN-LDA) scores. Darker colors indicate higher FUN-LDA scores, highlighting the functional significance of these variants in different tissues and cell types.

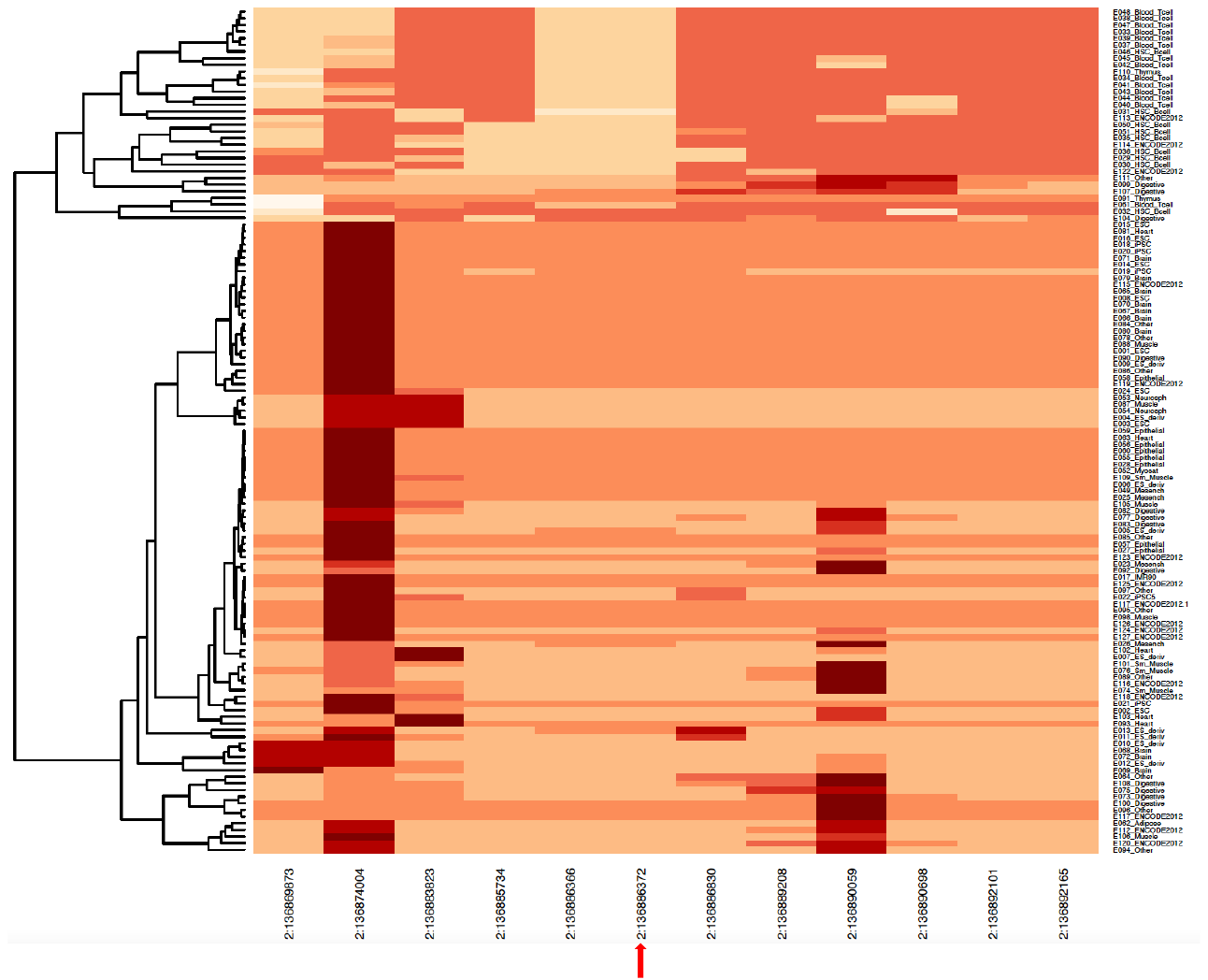

**Supplementary Figure 6:** Tissue-specific functional predictions were conducted for variants. The arrow indicates the index SNP (2:136886830). Additional columns along the X-axis represent all SNP proxies for the index variant, defined by having an R^2^ value of at least 0.8. These SNPs were arranged based on their genomic positions. The 127 tissues and cell types from the Roadmap Epigenomics project were clustered along the Y-axis using a hierarchical clustering approach. This clustering was based on the pattern of their functional latent Dirichlet allocation (FUN-LDA) scores. Darker colors indicate higher FUN-LDA scores, highlighting the functional significance of these variants in different tissues and cell types.

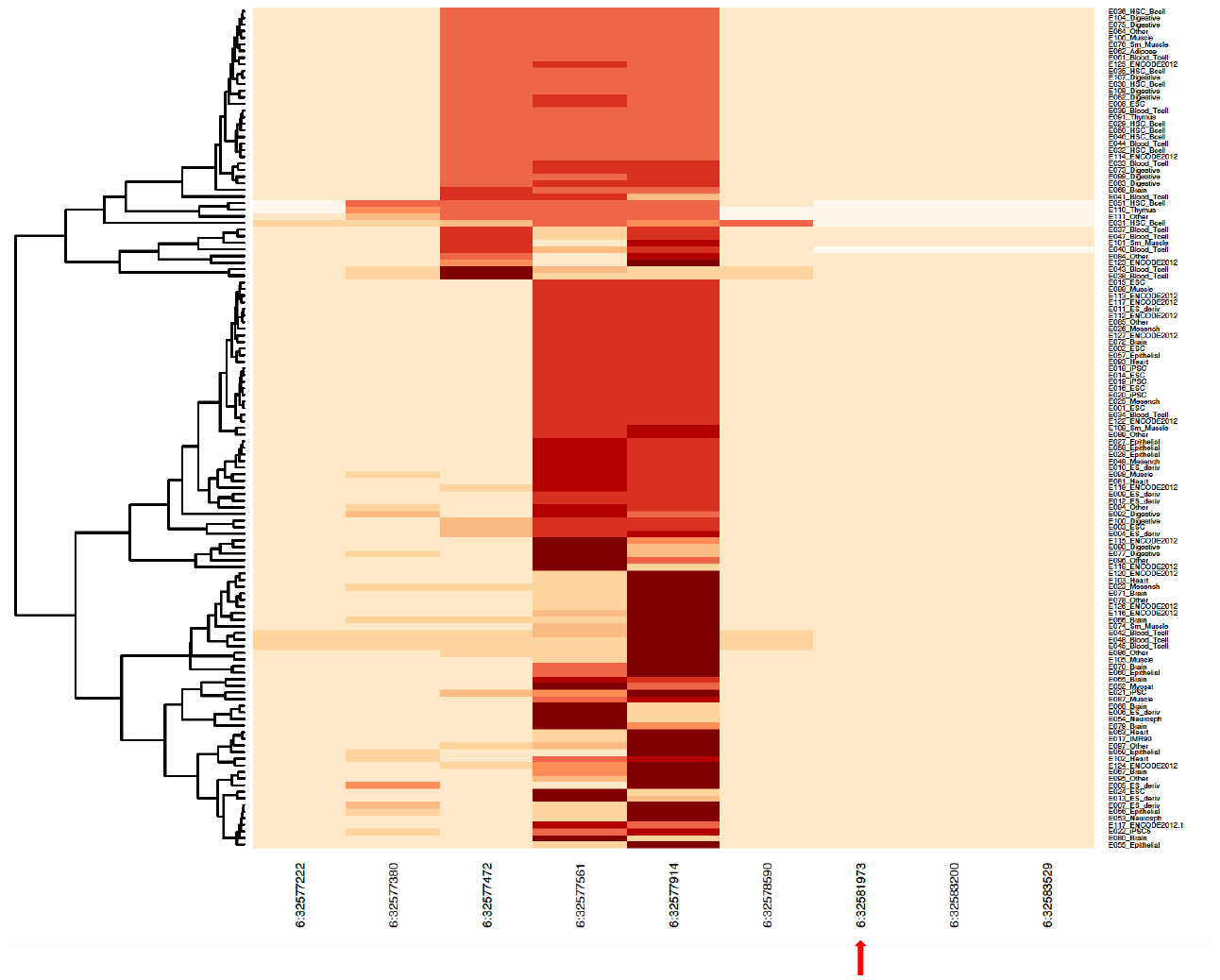

**Supplementary Figure 7:** Tissue-specific functional predictions were conducted for variants. The arrow indicates the index SNP (6:32581973). Additional columns along the X-axis represent all SNP proxies for the index variant, defined by having an R^2^ value of at least 0.8. These SNPs were arranged based on their genomic positions. The 127 tissues and cell types from the Roadmap Epigenomics project were clustered along the Y-axis using a hierarchical clustering approach. This clustering was based on the pattern of their functional latent Dirichlet allocation (FUN-LDA) scores. Darker colors indicate higher FUN-LDA scores, highlighting the functional significance of these variants in different tissues and cell types.

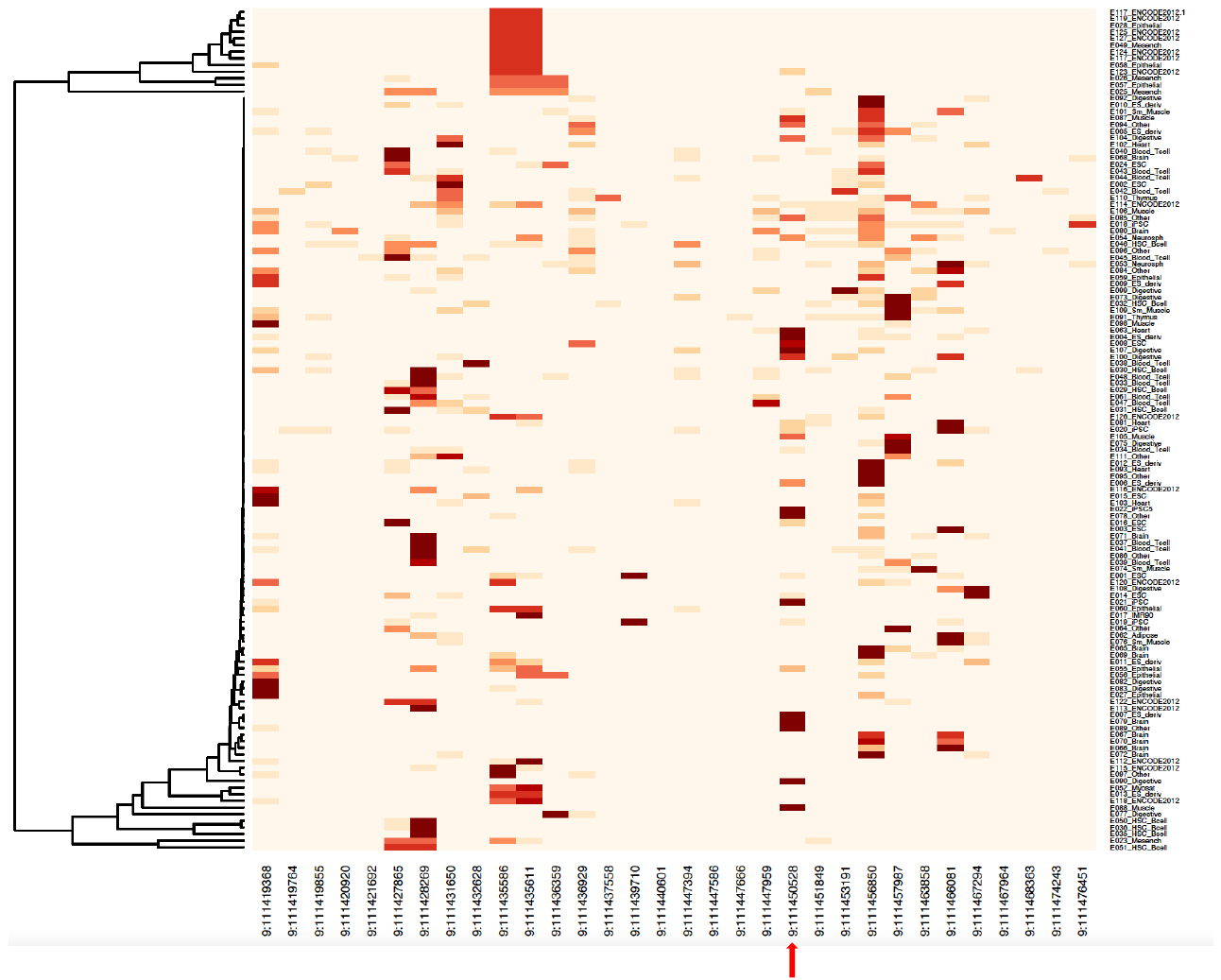

**Supplementary Figure 8:** Tissue-specific functional predictions were conducted for variants. The arrow indicates the index SNP (9:111450528). Additional columns along the X-axis represent all SNP proxies for the index variant, defined by having an R^2^ value of at least 0.8. These SNPs were arranged based on their genomic positions. The 127 tissues and cell types from the Roadmap Epigenomics project were clustered along the Y-axis using a hierarchical clustering approach. This clustering was based on the pattern of their functional latent Dirichlet allocation (FUN-LDA) scores. Darker colors indicate higher FUN-LDA scores, highlighting the functional significance of these variants in different tissues and cell types.

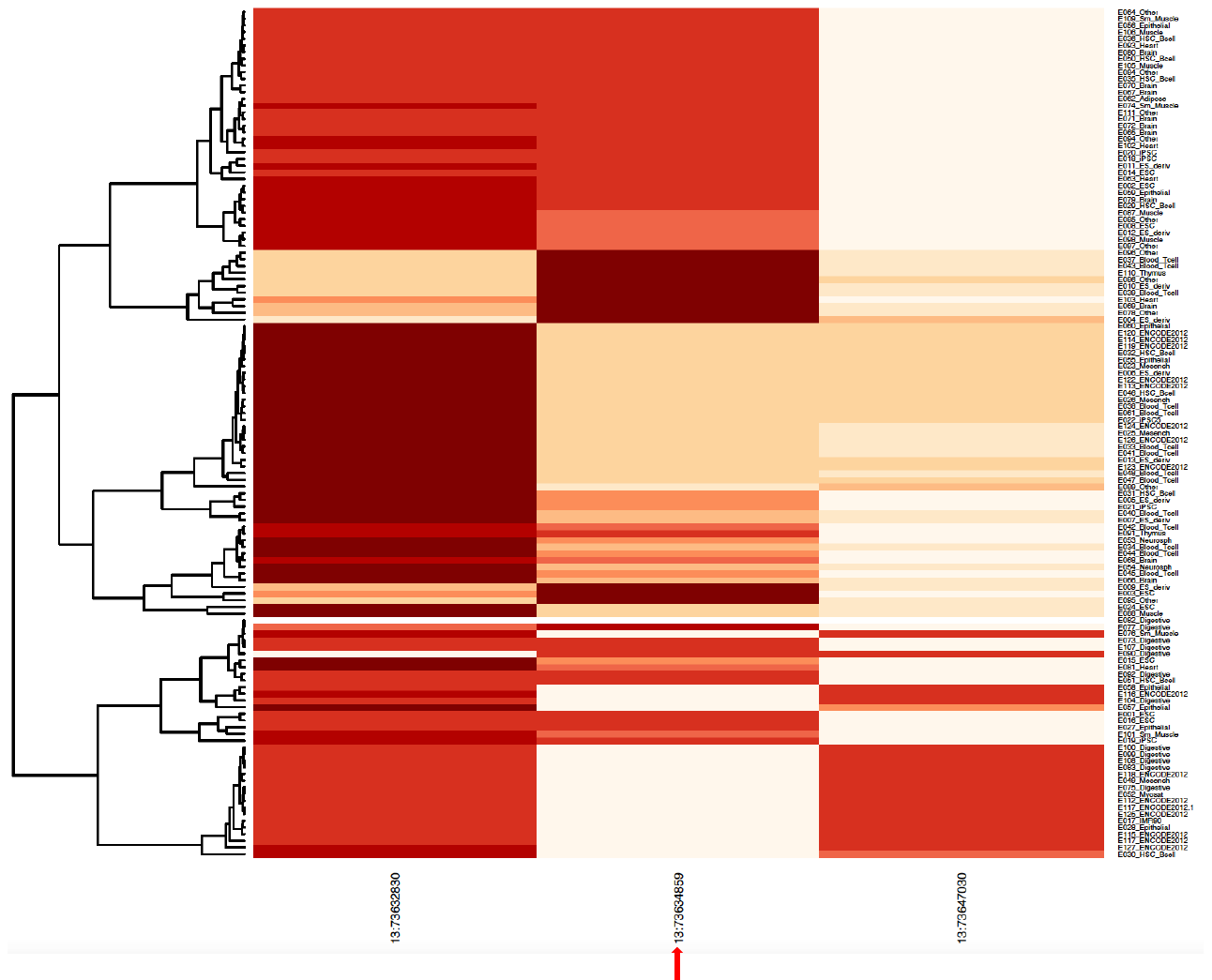

**Supplementary Figure 9:** Tissue-specific functional predictions were conducted for variants. The arrow indicates the index SNP (13:73634859). Additional columns along the X-axis represent all SNP proxies for the index variant, defined by having an R^2^ value of at least 0.8. These SNPs were arranged based on their genomic positions. The 127 tissues and cell types from the Roadmap Epigenomics project were clustered along the Y-axis using a hierarchical clustering approach. This clustering was based on the pattern of their functional latent Dirichlet allocation (FUN-LDA) scores. Darker colors indicate higher FUN-LDA scores, highlighting the functional significance of these variants in different tissues and cell types.

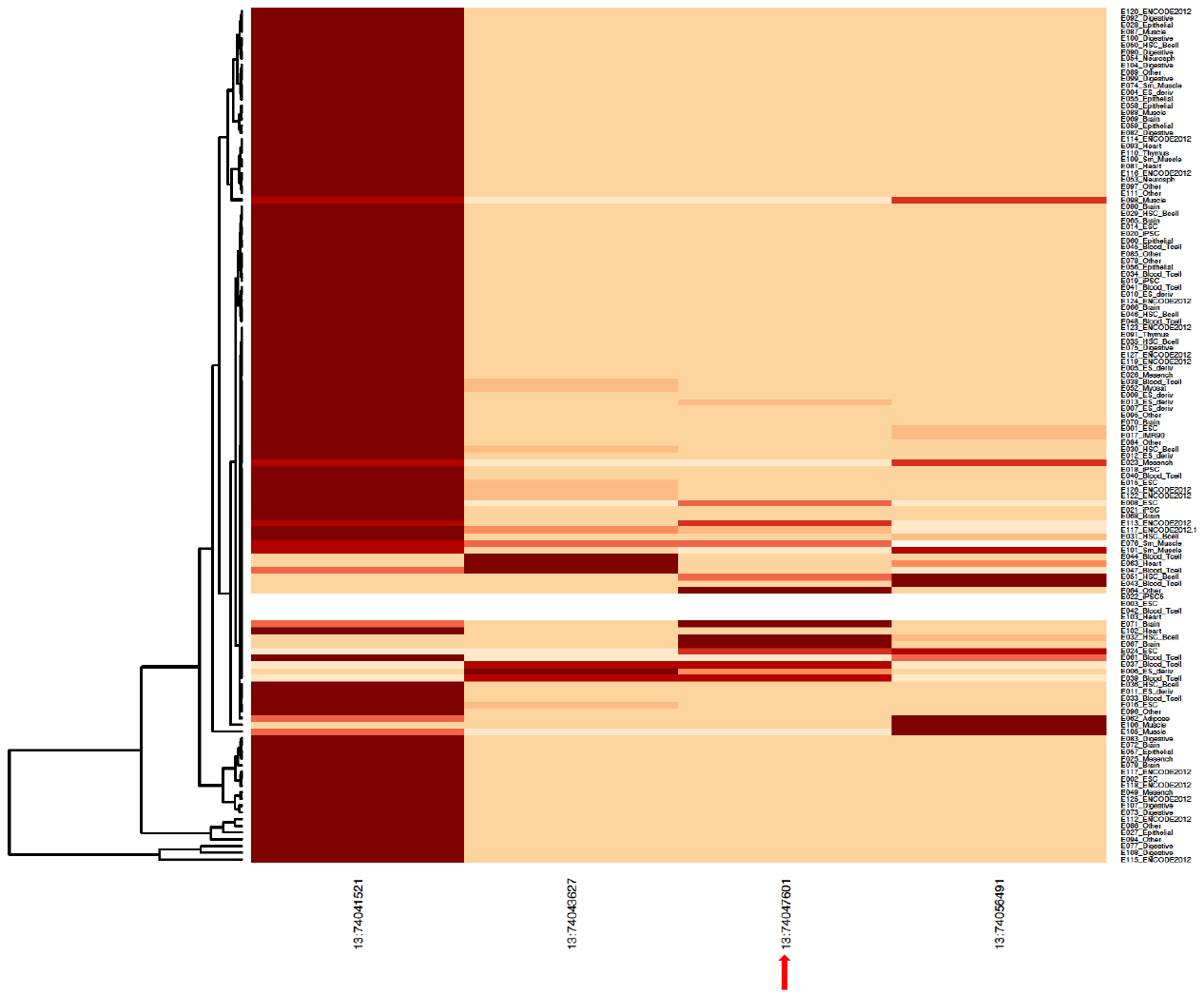

**Supplementary Figure 10:** Tissue-specific functional predictions were conducted for variants. The index SNP (13:74047601) is indicated by the arrow. Additional columns along the X-axis represent all SNP proxies for the index variant, defined by having an R^2^ value of at least 0.8. These SNPs were arranged based on their genomic positions. The 127 tissues and cell types from the Roadmap Epigenomics project were clustered along the Y-axis using a hierarchical clustering approach. This clustering was based on the pattern of their functional latent Dirichlet allocation (FUN-LDA) scores. Darker colors indicate higher FUN-LDA scores, highlighting the functional significance of these variants in different tissues and cell types.

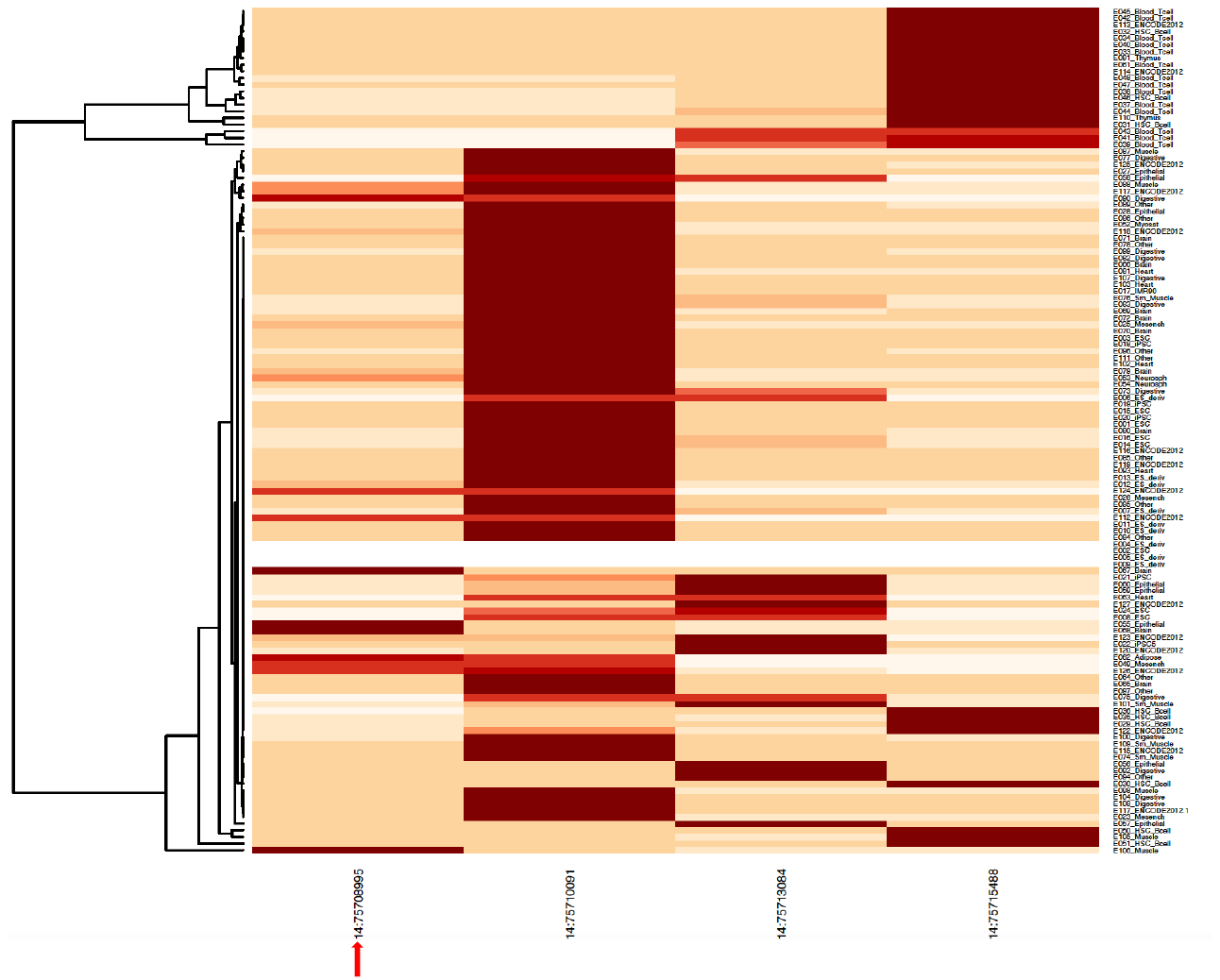

**Supplementary Figure 11:** Tissue-specific functional predictions were conducted for variants. The arrow indicates the index SNP (14:75709995). Additional columns along the X-axis represent all SNP proxies for the index variant, defined by having an R^2^ value of at least 0.8. These SNPs were arranged based on their genomic positions. The 127 tissues and cell types from the Roadmap Epigenomics project were clustered along the Y-axis using a hierarchical clustering approach. This clustering was based on the pattern of their functional latent Dirichlet allocation (FUN-LDA) scores. Darker colors indicate higher FUN-LDA scores, highlighting the functional significance of these variants in different tissues and cell types.

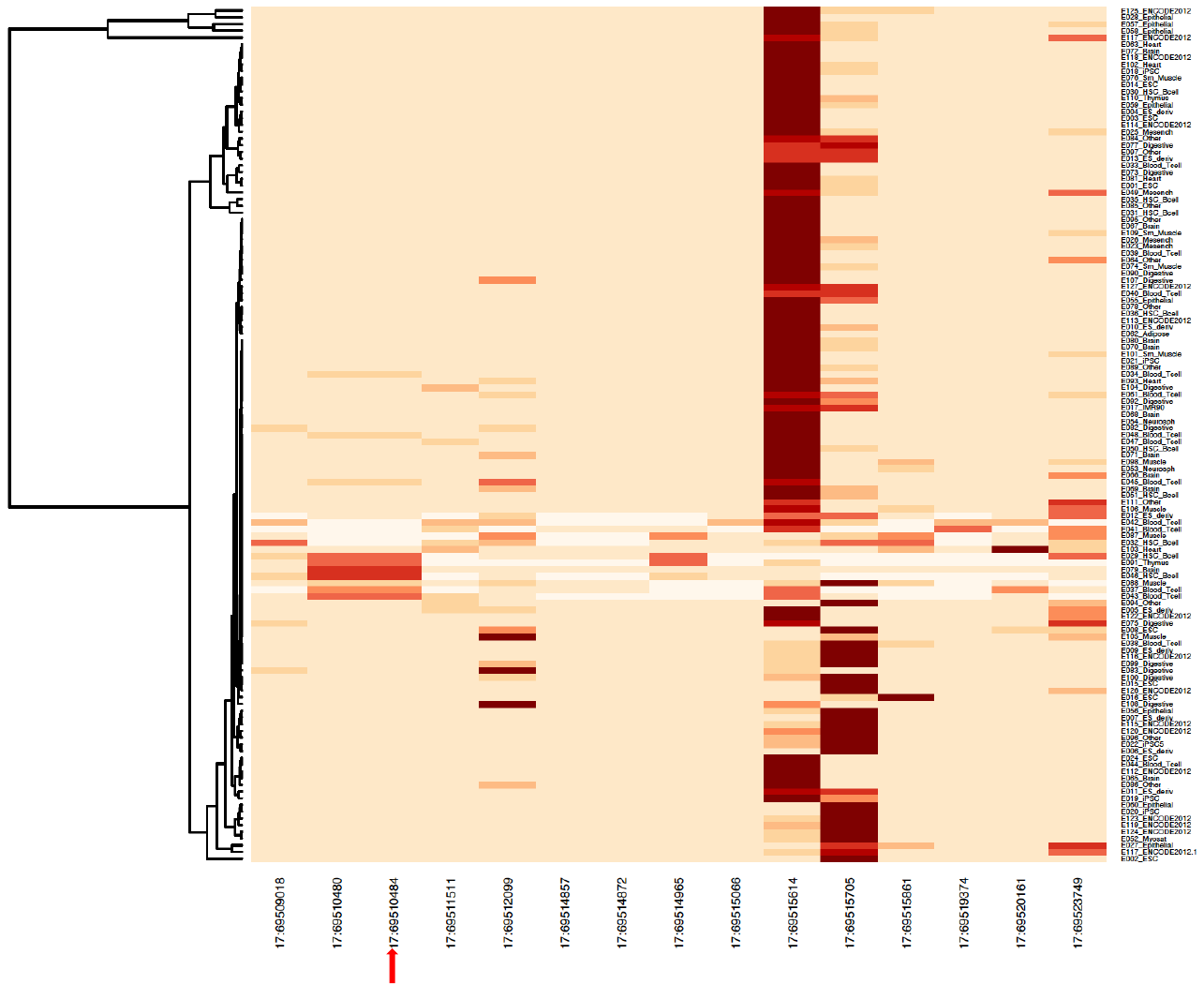

**Supplementary Figure 12:** Tissue-specific functional predictions were conducted for variants. The arrow indicates the index SNP (17:69510484). Additional columns along the X-axis represent all SNP proxies for the index variant, defined by having an R^2^ value of at least 0.8. These SNPs were arranged based on their genomic positions. The 127 tissues and cell types from the Roadmap Epigenomics project were clustered along the Y-axis using a hierarchical clustering approach. This clustering was based on the pattern of their functional latent Dirichlet allocation (FUN-LDA) scores. Darker colors indicate higher FUN-LDA scores, highlighting the functional significance of these variants in different tissues and cell types.

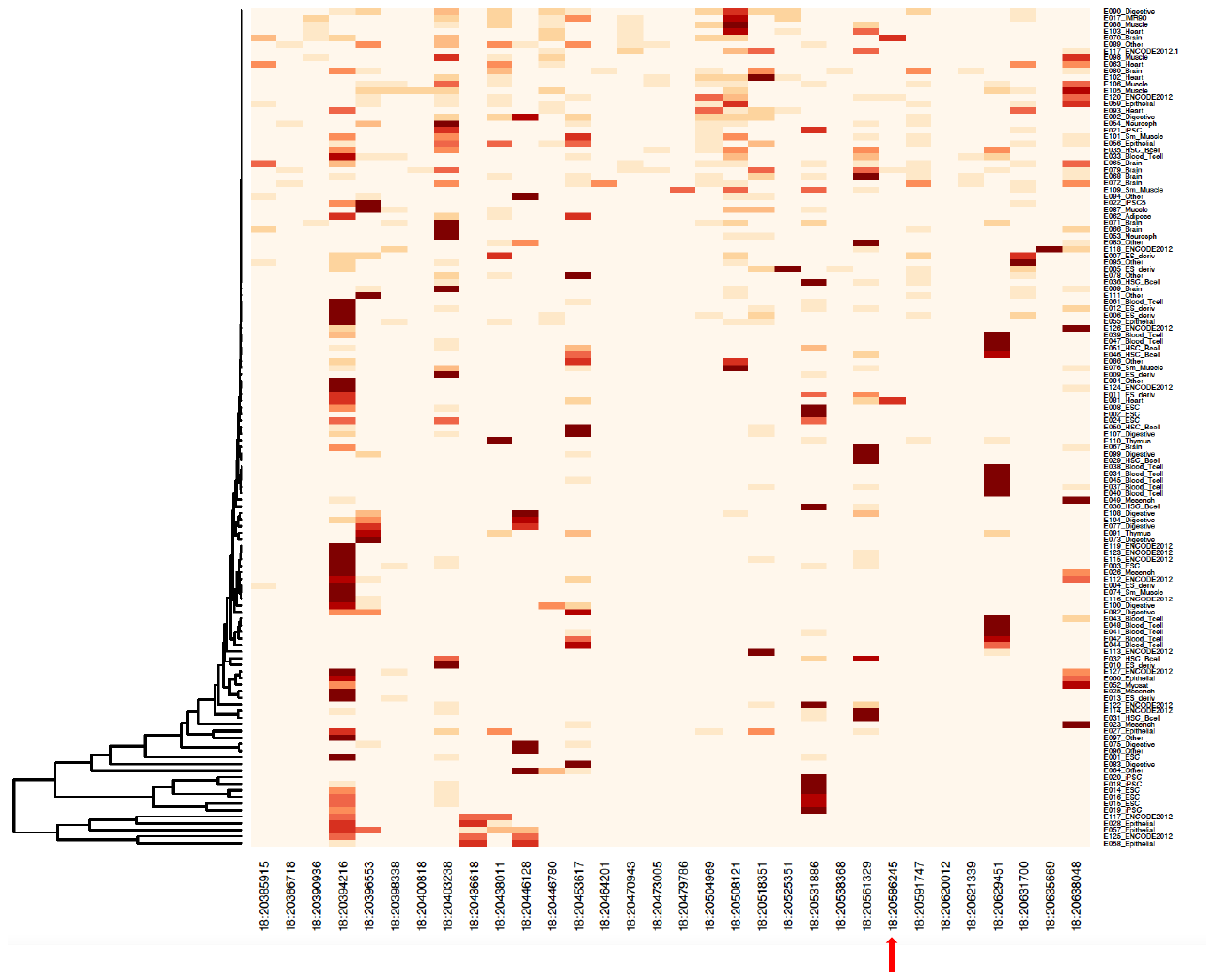

**Supplementary Figure 13:** Tissue-specific functional predictions were conducted for variants. The arrow indicates the index SNP (18:20586245). Additional columns along the X-axis represent all SNP proxies for the index variant, defined by having an R^2^ value of at least 0.8. These SNPs were arranged based on their genomic positions. The 127 tissues and cell types from the Roadmap Epigenomics project were clustered along the Y-axis using a hierarchical clustering approach. This clustering was based on the pattern of their functional latent Dirichlet allocation (FUN-LDA) scores. Darker colors indicate higher FUN-LDA scores, highlighting the functional significance of these variants in different tissues and cell types.

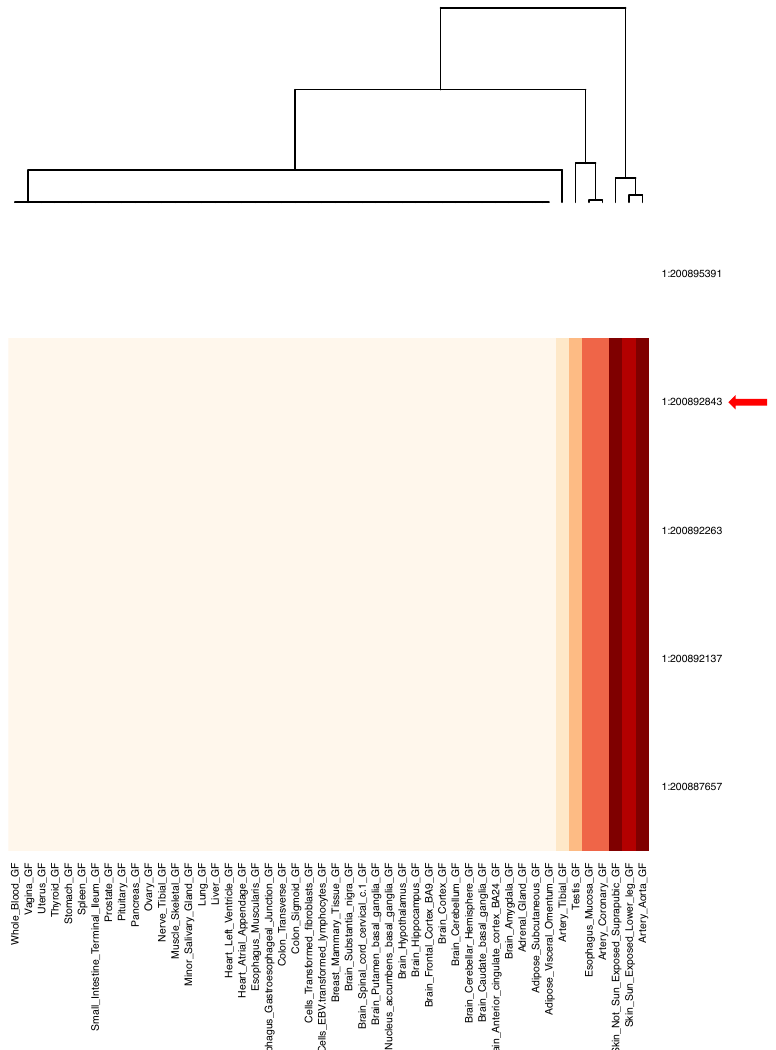

**Supplementary Figure 14:** Integration of variants with GTEx dataset. The arrow indicates the index SNP (1:200892843). Additional columns along the y-axis represent all SNP proxies for the index variant, defined by having an R2 value of at least 0.8. These SNPs were arranged based on their genomic positions. The 48 tissues from GTEx were clustered along the X-axis using a hierarchical clustering approach. This clustering was based on the P-values from the GTEx dataset. Darker colors indicate lower GTEx tissue Q-values, highlighting the significance of these variants in different tissues from GTEx.

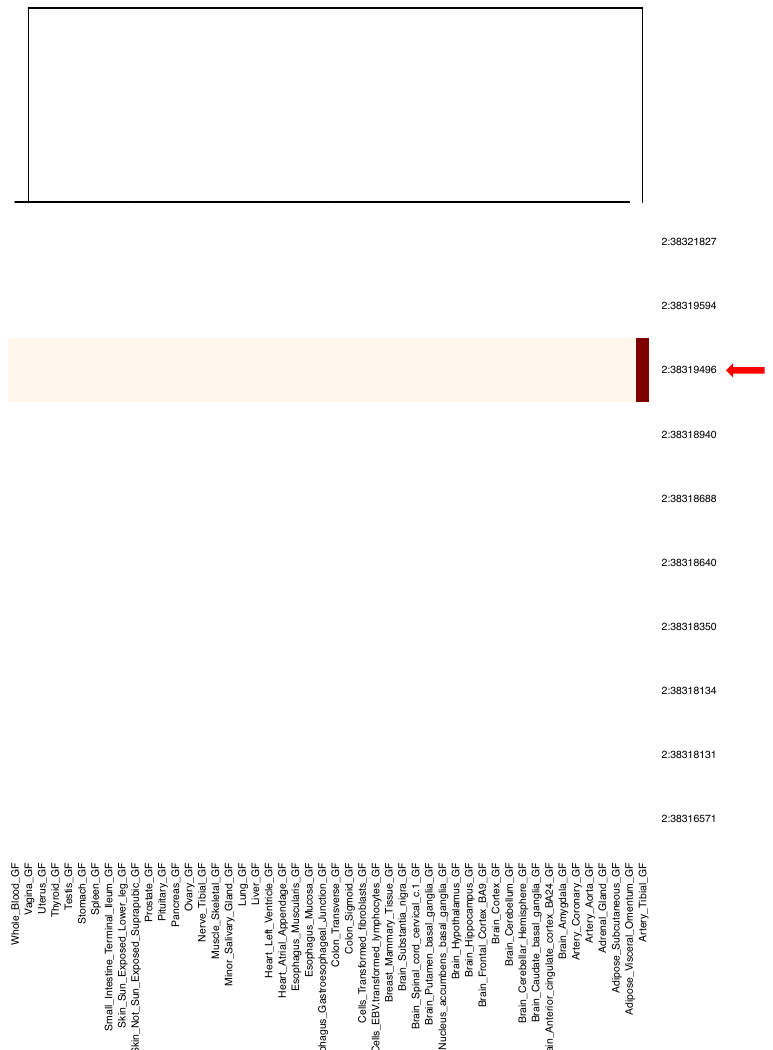

**Supplementary Figure 15:** Integration of variants with GTEx dataset. The arrow indicates the index SNP (2:38319496). Additional columns along the y-axis represent all SNP proxies for the index variant, defined by having an R2 value of at least 0.8. These SNPs were arranged based on their genomic positions. The 48 tissues from GTEx were clustered along the X-axis using a hierarchical clustering approach. This clustering was based on the P-values from the GTEx dataset. Darker colors indicate lower GTEx tissue Q-values, highlighting the significance of these variants in different tissues from GTEx.

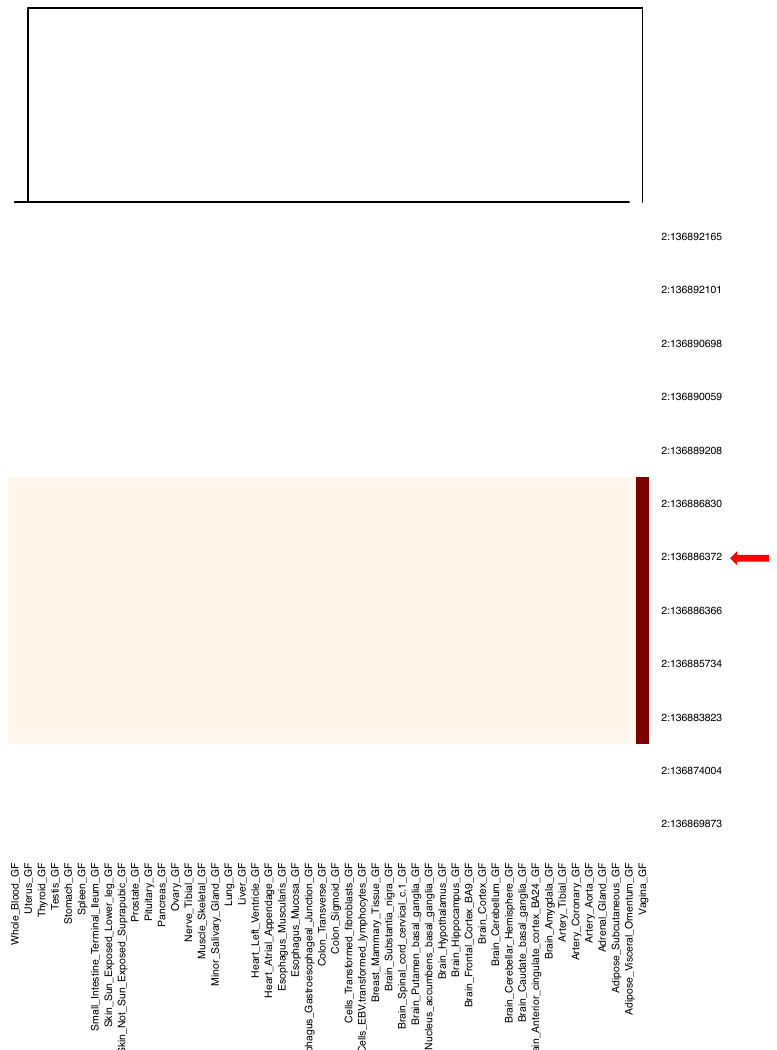

**Supplementary Figure 16:** Integration of variants with GTEx dataset. The arrow indicates the index SNP (2:136886372). Additional columns along the y-axis represent all SNP proxies for the index variant, defined by having an R2 value of at least 0.8. These SNPs were arranged based on their genomic positions. The 48 tissues from GTEx were clustered along the X-axis using a hierarchical clustering approach. This clustering was based on the P-values from the GTEx dataset. Darker colors indicate lower GTEx tissue Q-values, highlighting the significant of these variants in different tissues from GTEx.

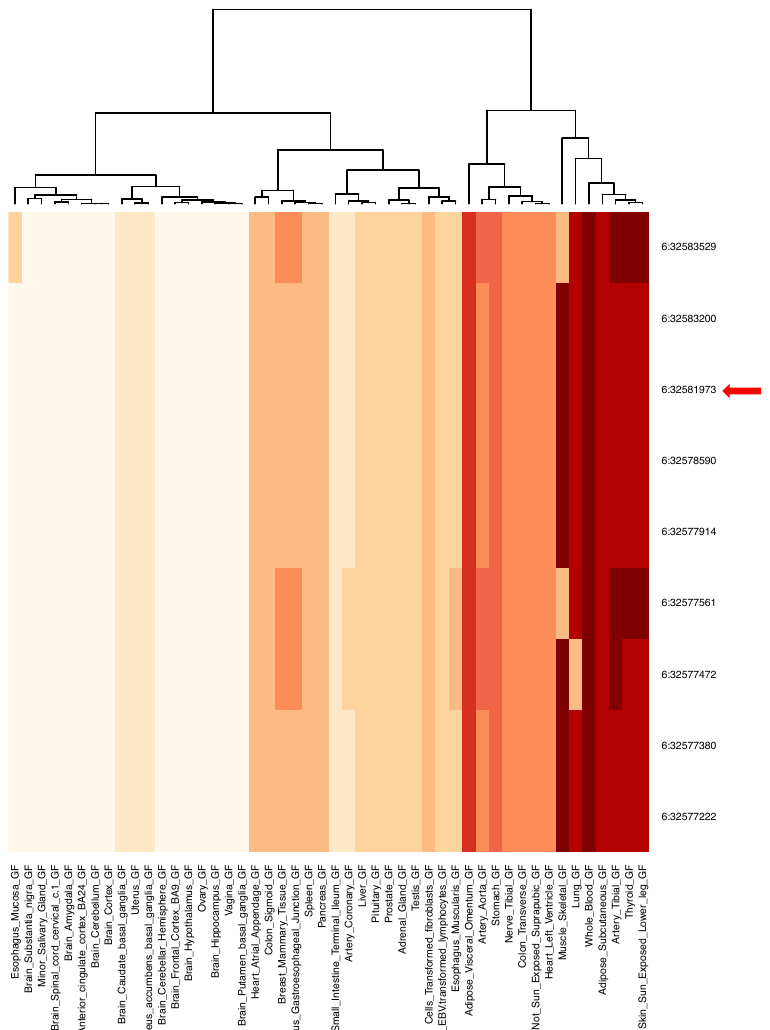

**Supplementary Figure 17:** Integration of variants with GTEx dataset. The index SNP (6:32581973) is indicated by the arrow. Additional columns along the y-axis represent all SNP proxies for the index variant, defined by having an R2 value of at least 0.8. These SNPs were arranged based on their genomic positions. The 48 tissues from GTEx were clustered along the X-axis using a hierarchical clustering approach. This clustering was based on the P-values from the GTEx dataset. Darker colors indicate lower GTEx tissue Q-values, highlighting the significant of these variants in different tissues from GTEx.

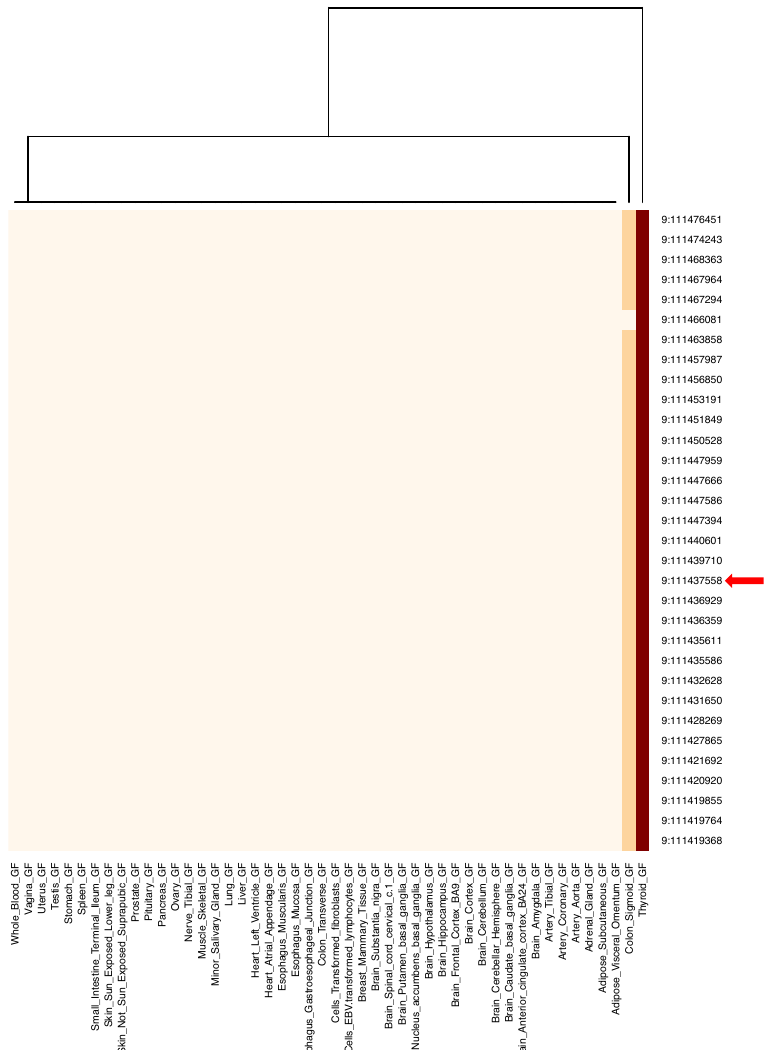

**Supplementary Figure 18:** Integration of variants with GTEx dataset. The index SNP (9:111437558) is indicated by the arrow. Additional columns along the y-axis represent all SNP proxies for the index variant, defined by having an R2 value of at least 0.8. These SNPs were arranged based on their genomic positions. The 48 tissues from GTEx were clustered along the X-axis using a hierarchical clustering approach. This clustering was based on the P-values from the GTEx dataset. Darker colors indicate lower GTEx tissue Q-values, highlighting the significant of these variants in different tissues from GTEx.

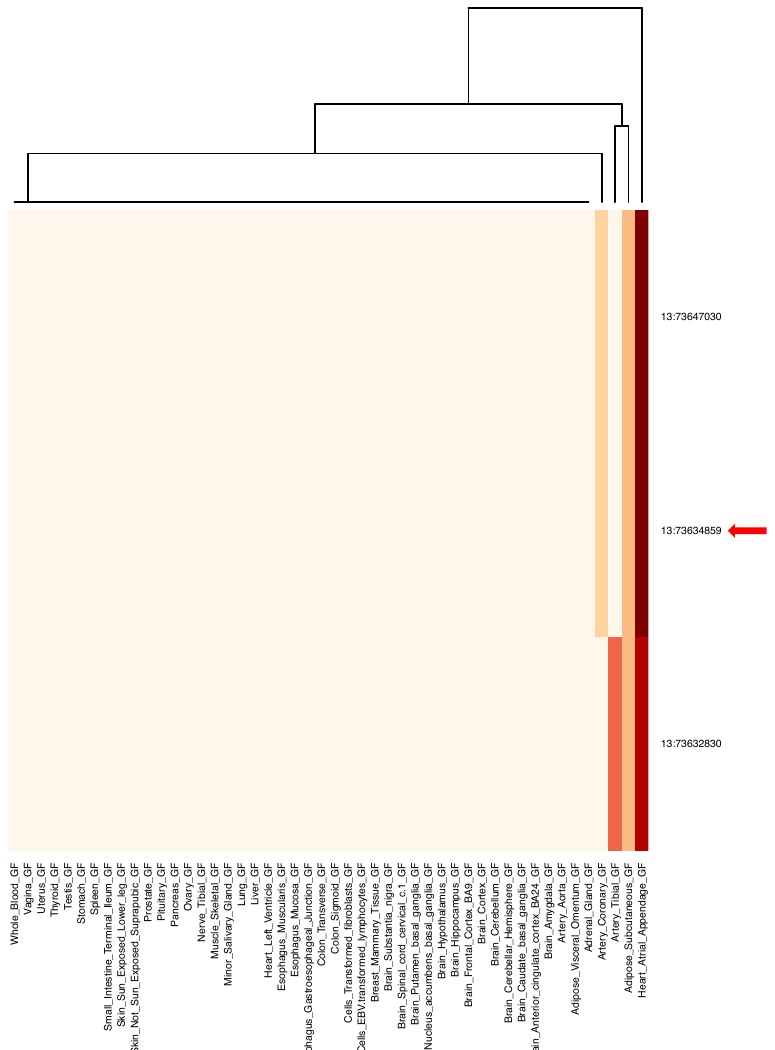

**Supplementary Figure 19:** Integration of variants with GTEx dataset. The arrow indicates the index SNP (13:73634859). Additional columns along the y-axis represent all SNP proxies for the index variant, defined by having an R2 value of at least 0.8. These SNPs were arranged based on their genomic positions. The 48 tissues from GTEx were clustered along the X-axis using a hierarchical clustering approach. This clustering was based on the P-values from the GTEx dataset. Darker colors indicate lower GTEx tissue Q-values, highlighting the significance of these variants in different tissues from GTEx.

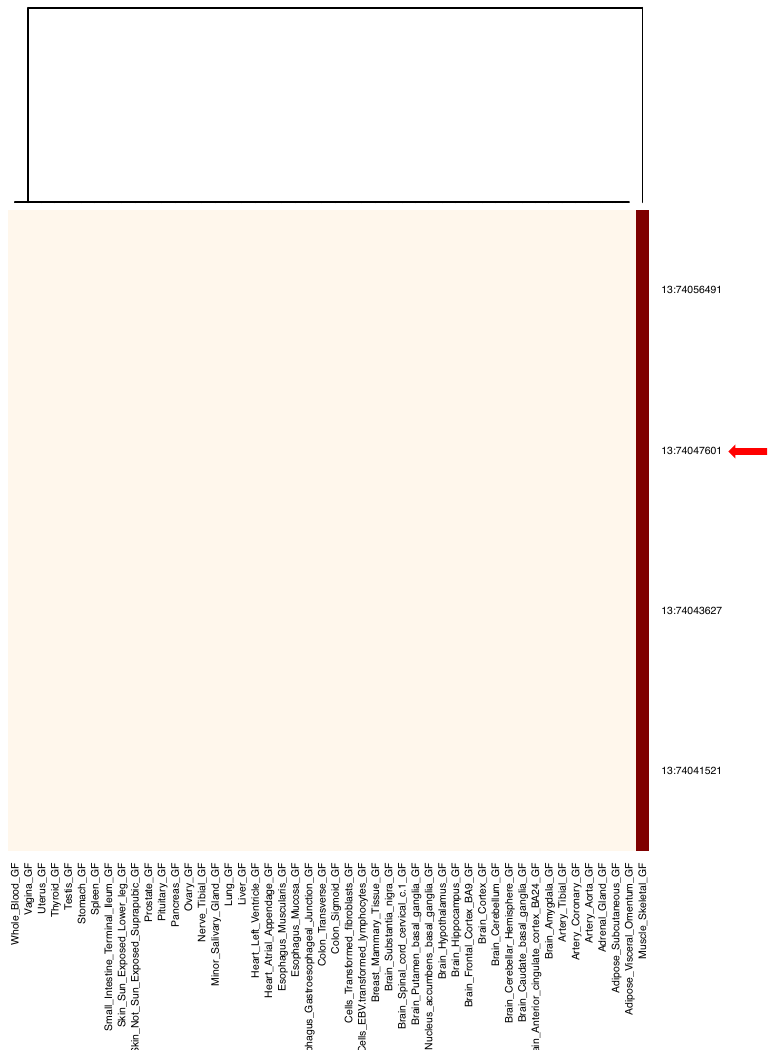

**Supplementary Figure 20:** Integration of variants with GTEx dataset. The arrow indicates the index SNP (13:74047601). Additional columns along the y-axis represent all SNP proxies for the index variant, defined by having an R2 value of at least 0.8. These SNPs were arranged based on their genomic positions. The 48 tissues from GTEx were clustered along the X-axis using a hierarchical clustering approach. This clustering was based on the P-values from the GTEx dataset. Darker colors indicate lower GTEx tissue Q-values, highlighting the significance of these variants in different tissues from GTEx.

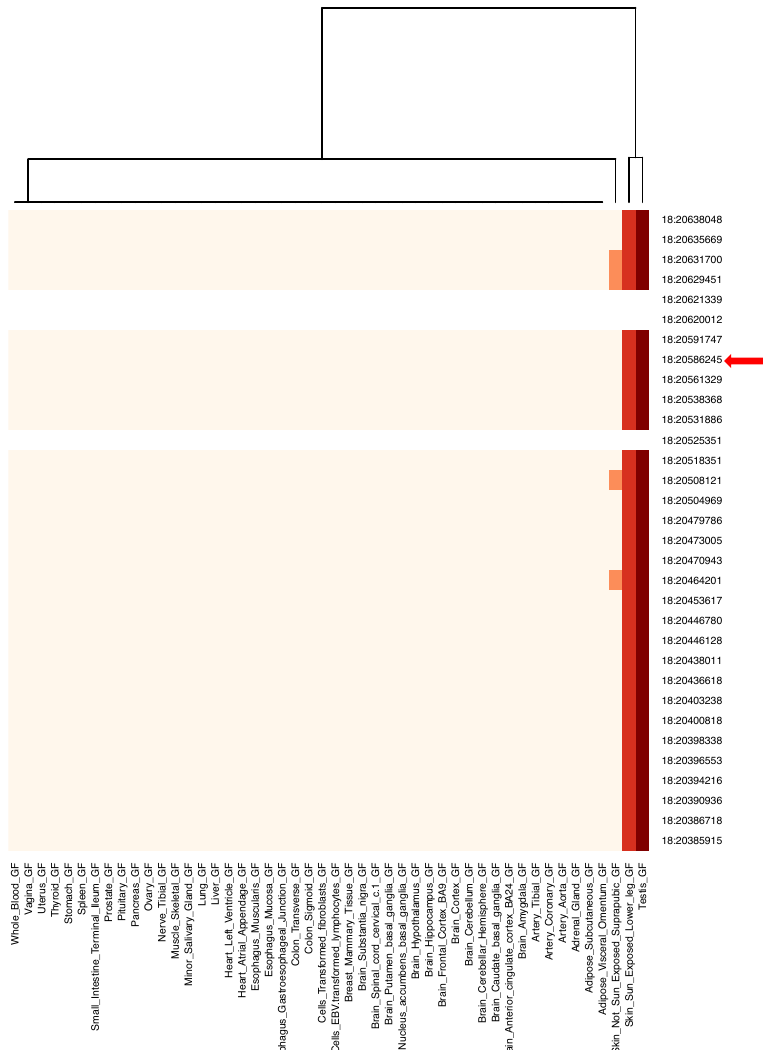

**Supplementary Figure 21:** Integration of variants with GTEx dataset. The arrow indicates the index SNP (18:20586245). Additional columns along the y-axis represent all SNP proxies for the index variant, defined by having an R2 value of at least 0.8. These SNPs were arranged based on their genomic positions. The 48 tissues from GTEx were clustered along the X-axis using a hierarchical clustering approach. This clustering was based on the P-values from the GTEx dataset. Darker colors indicate lower GTEx tissue Q-values, highlighting the significance of these variants in different tissues from GTEx.

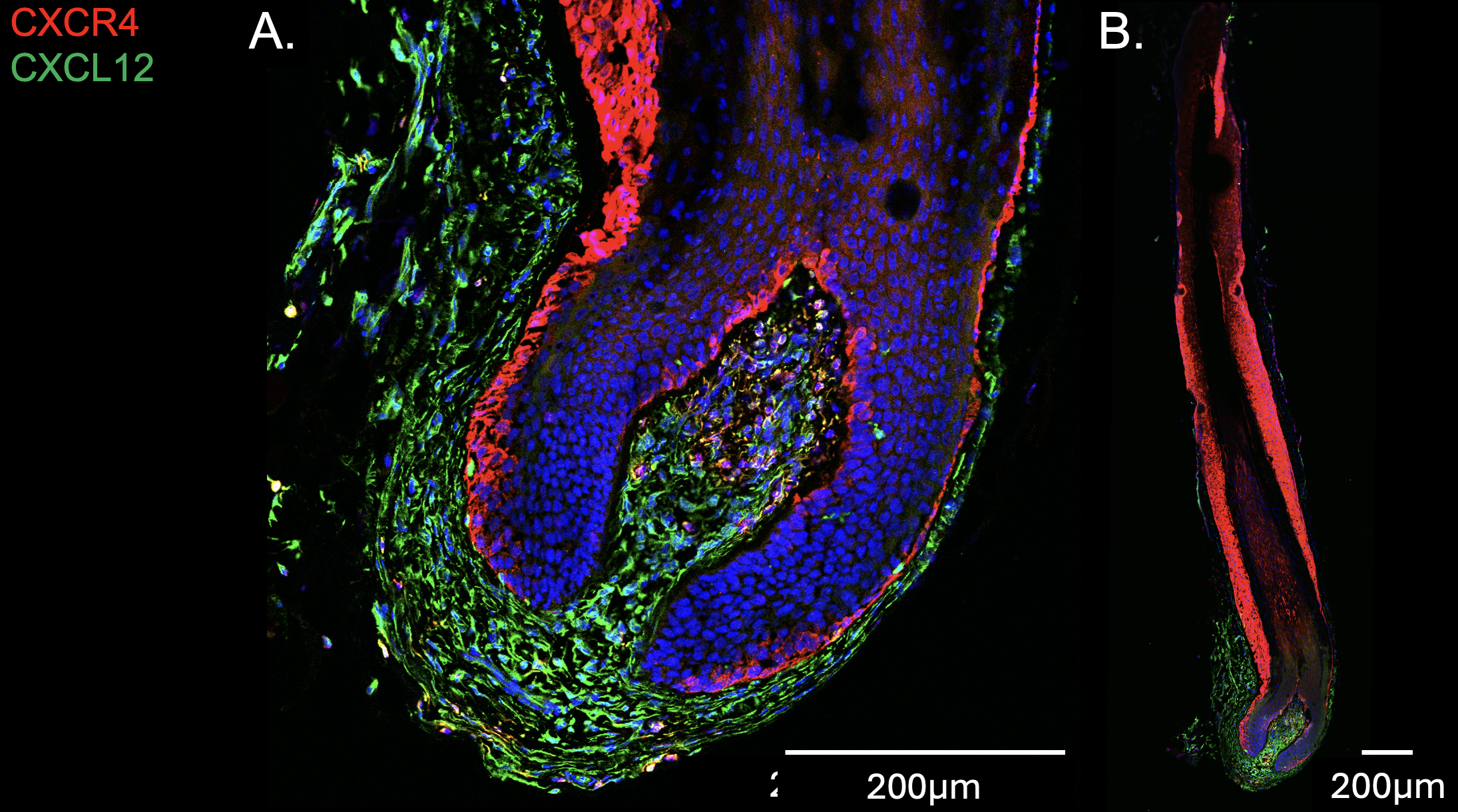

**Supplementary Figure 22. Characterization of CXCR4 and CXCL12 expression in human terminal hair** **follicles**. A. and B. CXCR4 expression (red) localizes to the outer root sheath and a few dermal papilla cells in human beard hair follicles. CXCL12 expression is found in both the dermal sheath and dermal papilla.

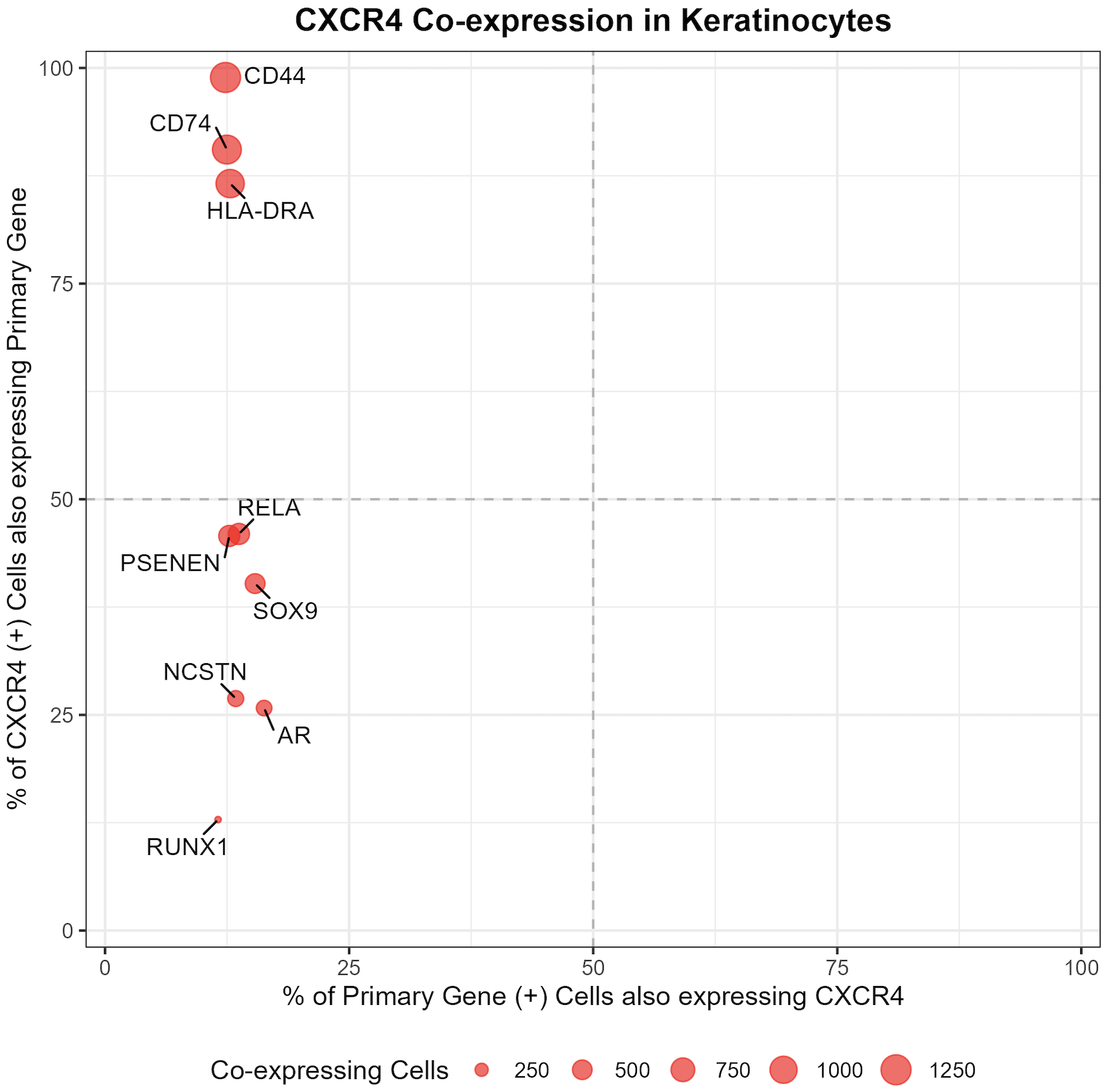

**Supplementary Figure 23. Genes co-expressed in *CXCR4* HS Keratinocytes.** Single cell RNA-sequencing in HS skin (GSE154775) was used to identify genes relevant to Figure 2D. The Y-axis represents the percentage of CXCR4+ keratinocytes that also express a gene of interest. For example, HLA class II genes implicated in our GWAS are also highly co-expressed. Furthermore, 100% of HS keratinocytes expressing *CXCR4* also express *CD44*. Both of these genes are co-receptors for CD74 (*30*). *NCSTN* and *PSENEN* are γ-secretase genes implicated in HS pathogenesis by rare pathogenic variants and common risk alleles (*31, 32*). The γ-secretase complex cleaves the intracellular domain of CD74 releasing it into the cytoplasm, where it can form complexes with RELA or RUNX1 and subsequently translocate to the nucleus to activate NF-κB genes (*33, 34*). The androgen receptor (AR) is upregulated in HS CXCR4+ keratinocytes relative to CXCR4- keratinocytes. The AR has previously been implicated in HS pathogenesis in transcriptomic studies, which showed expression in epithelial cells of the epidermis and epithelialized epidermal tunnels (*35*).

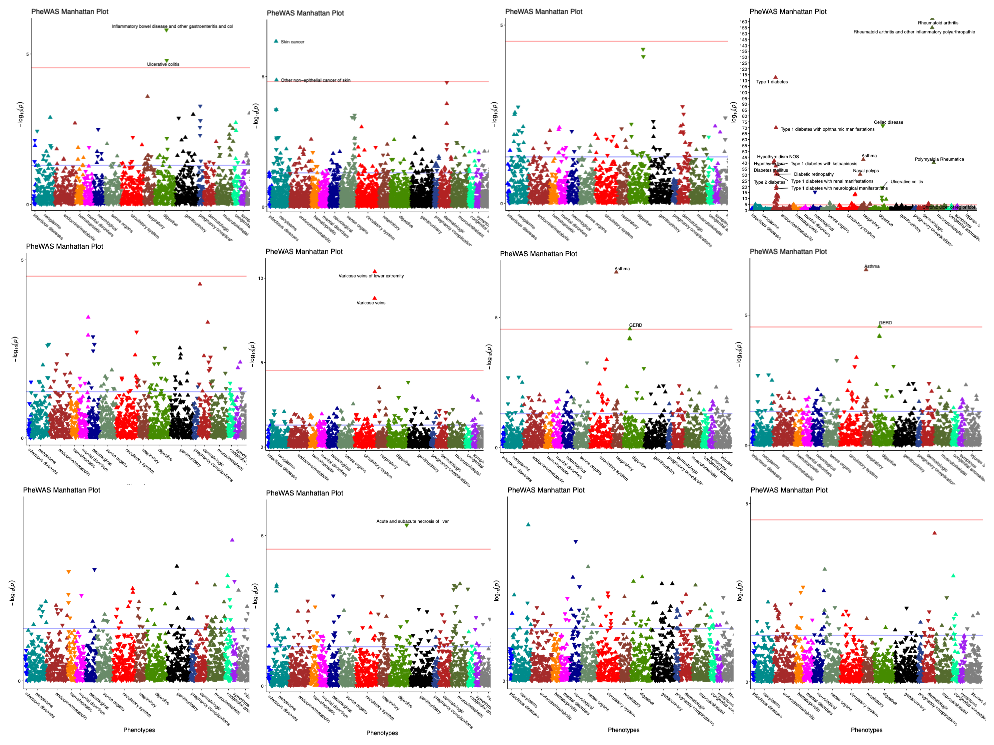

**Supplementary Figure 24:** We conducted PheWAS for each independent genome-wide significant locus and identified pleiotropic associations at five loci, identifying HS risk alleles shared with asthma, skin cancer, actinic keratosis varicose veins, and multiple immune-mediated diseases at the HLA (upward triangles). Negative associations implying opposing effects were identified for inflammatory bowel disease, ulcerative colitis, Celiac disease, and pain, among others.
